# A behavioural modelling approach to assess the impact of COVID-19 vaccine hesitancy

**DOI:** 10.1101/2021.06.22.21259324

**Authors:** Bruno Buonomo, Rossella Della Marca, Alberto d’Onofrio, Maria Groppi

## Abstract

In this paper we introduce a compartmental epidemic model describing the transmission of the COVID-19 disease in presence of non-mandatory vaccination. The model takes into account the hesitancy and refusal of vaccination. To this aim, we employ the information index, which mimics the idea that individuals take their decision on vaccination based not only on the present but also on the past information about the spread of the disease. Theoretical analysis and simulations show clearly as a voluntary vaccination can certainly reduce the impact of the disease but it is unable to eliminate it. We also show how the information-related parameters affect the dynamics of the disease. In particular, the hesitancy and refusal of vaccination is better contained in case of large information coverage and small memory characteristic time. Finally, the possible influence of seasonality is also investigated.

## 1 Introduction

On 31 December 2019, the Chinese public health authorities reported to WHO the existence in Wuhan City of a cluster of cases of viral pneumonia [73]. The causal agent of the disease was shortly later identified as a new type of SARS, and named SARS-CoV-2. Although many governments undervalued the pandemic risks [55], since 21 January 2020 WHO published on its website a daily situation reports. In the first report it is clearly written *WHO has issued interim guidance for countries, updated to take into account the current situation* [73]. Indeed, the first extra-China case was on 13 January 2020 and then rapidly moved in other countries. Finally, it developed in a devastating pandemics we all know, causing the temporary collapse of many health systems. For example, France in the pre COVID-19 era had about 5000 ICU beds, however at the peak of its first wave 7019 ICU beds were occupied by COVID-19 patients [32].

In the first year of the pandemic, in the absence of a vaccine, the only possible pandemic mitigation strategies were locally based on social distancing and partial and full lockdowns [8, 55]. Lockdowns were generally very effective in reducing the pressure of the pandemic on the health systems of the countries but the period after them was generally characterized by a new epidemic outbreak after some months. Up to now most countries had three epidemic outbreaks (also termed *waves*) [14].

Since the early stage of the pandemic, many authors implemented models, from traditional mathematical epidemiology, for the evolution and control of COVID-19 disease [16, 19, 22, 33, 34, 46, 63]. The early dynamics of transmission in Wuhan, China, was studied by Kucharski *et al*. [46] through a stochastic SEIR model using the data obtained from the outbreak in Wuhan. Gatto *et al*. [33] proposed a model to study the transmission between a network of 107 Italian provinces during the initial stage of the first COVID-19 wave. A network model applied to Italy was proposed also by Della Rossa *et al*. [19] to show that heterogeneity between regions plays a fundamental role in designing effective strategies to control the disease while preventing national lockdowns. Giordano *et al*. [34] introduced a model for assessing the effectiveness of testing and contact tracing combined with social distancing measures. Non-pharmaceutical interventions to fight COVID-19 in the UK and US were considered by Davis *et al*. [16] and Ngonghala *et al*. [63], respectively, while the effect of social distancing during lockdown in France was studied by Dolbeault and Turinici [22] by using a variant of the SEIR model. Many other relevant studies focused on assessing the effects of containment measures and predicting epidemic peaks and ICU accesses, see e.g. [29, 31, 68]. As soon as vaccines for COVID-19 became available, many compartmental models have began to appear in the literature with the specific aim of investigating the vaccination effects on the spread of the disease as well as assessing the optimal allocation of vaccine supply [5, 15, 20, 60].

A limitation of classical Mathematical Epidemiology (ME) is that it is built up on Statistical Mechanics: the agents are modelled as if they were molecules and the contagion is abstracted as a chemical reaction between ‘molecules’ of the healthy species with ‘molecules’ of the infectious species. Thus, mass action-like laws are used in such models. The missing ingredient of ME is the behaviour of agents: how people modify their contacts at risk and how their vaccine-related decisions are taken. The absence of this ingredient makes models of classical ME increasingly less adapt as a tool for Public Health. Indeed, a major challenge for global Public Health is the spread of hesitancy and refusal of vaccines. This is due to the phenomenon of ‘Pseudo-Rational’ Objection to VAccination (PROVA) [10]: people overweight real and imaginary side effects of vaccines and underweight real risks due to the target infectious diseases [10, 56, 72]. PROVA is inducing remarkable changes in the civil society attitude towards the prevention of infectious diseases. This increasingly important lack of trust towards vaccination is one of the many negative consequences of two distinct and synergyzing phenomena of more general nature: the *post-trust society* [51] and the *post-truth era* [58].

The first work that explicitly modelled social distancing in ME was [11], which incorporated a phenomenological behavioural response into the Kermack and McKendrick’s epidemic model. The emergence of PROVA led in the last two decades to the birth of a new branch of ME: the Behavioural Epidemiology of infectious diseases (BEID) [56,72]. The main aim of BEID is to embed the impact of human behaviour in models of the spread and control of infectious diseases [56, 72]. The key role of both present and past information on vaccination decisions and uptake as well as on the social distancing was first stressed, respectively, in [26,56] and in [23] by means of phenomenological models. In a recent paper [8] a model for the transmission of COVID-19 disease has been introduced. The model considers the social distancing and quarantine as mitigation strategies by the Public Health System. The model is information-dependent, in the sense that contact rate and quarantine rate are assumed to depend on the available information and rumours about the disease status in the community. In [8] the model is applied to the case of the COVID-19 epidemic in Italy. The paper estimates that citizen compliance with mitigation measures played a decisive role in curbing the epidemic curve, by preventing a duplication of deaths and about 46% more infections.

The COVID-19 pandemic caused a worldwide effort on the vaccine that resulted in the rapid development of new vaccines [45, 52], some of which belongs to the new class of mRNA vaccines [2, 64]. In the light of the deep changes in the life of milliards of people and of the huge negative impact on world economics that the world has experienced, one could have expected that only a tiny proportion of people would really be hesitant towards vaccination. Unfortunately, this is not what occurred. As early as June 2020 Neumann-Böhme and coworkers [62] investigated the attitudes about anti COVID-19 vaccination of a representative sample of citizens of seven European countries. Amazingly, although the first European epidemic wave had just ended, a large proportion of hesitancy and opposition to the vaccines were found in all class ages, and in both sex. In particular, in France the 38% of citizens were hesitant (28%) or strongly against (10%) anti COVID-19 vaccines.

Before mid December 2020 phase 3 of a number of vaccines ended, showing that they have a very outstanding effectiveness in preventing COVID-19 [2, 52, 64]. Typically, drug regulatory agencies defined priority groups for the vaccination (elderly people with serious co-morbidities, healthcare workers in senior residences, etc.). From a rational viewpoint there were all the premises to believe that the vaccine hesitancy would have been strongly reduced and that mandatory vaccination campaigns could have been conducted but this was not the case. As far as the mandatory nature of the vaccination campaign is concerned, in many countries the vaccines are no mandatory [48, 54, 69]. As for the vaccine hesitancy, an investigation conducted in October 2020 [38] suggests that 46% of French citizens are vaccine hesitant. Other countries have percentages of opposition and hesitancy that exceeds 30%: 36% in Spain and USA, 35% in Italy, 32% in South Africa, 31% in Japan and Germany. Globally, the hesitancy and objection area is as large as 27%.

Given these large percentages of hesitance and opposition to the COVID-19 vaccine, we think that applying the behavioural epidemiology approach to model the implementation of a vaccination campaign for COVID-19 is appropriate. To this end, we adopt a strategy remindful of the one used in [26]. Namely, we assume that the vaccination rate is a phenomenological function of the present and past information that the citizens have on the spread of the epidemic. Note that, in the context of SIR and SEIR infectious diseases, more mechanistic models based on evolutionary game theories [3, 24, 25, 72] exist, but reduce to the approach of [10, 26] in case of volatile opinion switching [18, 24, 72].

In this paper, we consider a COVID-19 affected population controlled by vaccination, where the final choice to vaccinate or not is partially determined on a fully voluntary basis and depends on the publicly available information on both present and recent past spreading of the disease in the community. Our model is inspired by the compartmental epidemic model introduced in [8], where the COVID-19 transmission during the 2020 lockdown in Italy was studied. In some sense, compared with the model in 8], the main difference is that here the non-pharmaceutical interventions (social distancing and quarantine) are replaced by vaccination. An analogous situation was considered by Gumel and co-workers for SARS epidemic in 2003 when they studied a SARS model in [37] and then considered vaccination intervention in [36].

We perform a qualitative analysis based on stability theory and bifurcation theory. The analysis shows that, when the control reproduction number, ℛ_*V*_, is less than 1, there exists only the disease-free equilibrium (DFE) that is globally stable; otherwise, when ℛ_*V*_ > 1, the DFE is unstable and an endemic equilibrium arises. The model is then parametrized based on the COVID-19 epidemic in Italy and on preliminary reports about anti COVID-19 vaccines. In numerical simulations, we consider two possible starting times for a one year-lasting vaccination campaign. We assess the role of vaccine and information-related parameters by evaluating how they affect suitable epidemiological indicators. Finally, the presence of seasonality effects is investigated by adding the assumption that the disease transmission and severity as well as the rate of vaccination, are lower during the warmer months.

The paper is organized as follows. In Section 2 the model is introduced and in Section 3 the qualitative analysis is performed. Model parametrization and numerical solutions are given in Section 4 and Section 5, respectively. The case of seasonally-varying parameter values is addressed in Section 6. Concluding remarks follow in Section 7. The paper is complemented by the Appendix A.

## 2 The model

### 2.1 State variables and the information index

We consider a population affected by COVID-19 disease, where a vaccine is available and administered on voluntary basis and not mandatory. We assume that the vaccine provides only partial protection, so that the transmission of the disease due to contacts between vaccinated and infectious individual is still possible, although with reduced probability. We also assume that both the vaccine-induced immunity and the disease-induced immunity are not waning (see Remark 1 below for a discussion on this point). The total population at time *t* (say, *N*) is divided into the following six disjoint *compartments*:

– *susceptibles, S*: individuals who are healthy but can contract the disease;
– *exposed* (or *latent*), *E*: individuals who are infected by SARS-CoV-2 but are not yet capable of transmitting the virus to others;
– *asymptomatic infectious, I*_*a*_: this compartment includes two groups, namely the *post-latent* individuals, i.e. individuals who lie in the phase of incubation period following latency, where they are infectious and asymptomatic, and the *truly asymptomatic* individuals, i.e. who have no symptoms throughout the course of the disease;
– *symptomatic infectious, I*_*s*_: infectious individuals who show mild or severe symptoms;
– *vaccinated, V*: individuals who are vaccinated with at least one dose of COVID-19 vaccine;
– *recovered, R*: individuals who are recovered after the infectious period.

The size of each compartment at time *t* represents a *state variable* of the mathematical model, and *N* = *S* + *E* + *I*_*a*_ + *I*_*s*_ + *V* + *R*.

We assume that agents take their decision on vaccination based not only on the present but also on the past information they have on the spread of the disease, the past being weighted in an exponential way. Therefore the information on the status of the disease in the community is described by means of the *information index* [26, 72]:

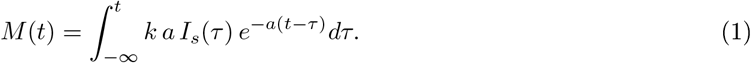

Such index is an important tool of behavioural epidemiology [56] and is an extension of the idea of the prevalence-dependent contact rate, developed by V. Capasso in the seventies, which describes the behavioural response of individuals to prevalence [11]. Here, the parameter *a* takes the meaning of inverse of the average time delay of the collected information on the disease (say, *T*_*a*_ = *a*^−1^) and the parameter *k* is the *information coverage*, which summarises two opposite phenomena: the disease under-reporting and the level of media coverage of the disease status, which tends to amplify the social alarm. It may be assumed that *k* ∈ (0, 1], see [9].

From (1), by applying the linear chain trick [53], we obtain the differential equation 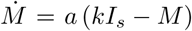, ruling the dynamics of *M*.

#### Remark 1.

*Together with the role of human behaviour in the vaccine decisions, the other major hypotheses of the above model are that the vaccine is not perfect and there is no waning effect of both natural and vaccine-induced immunity. The first is related to the scientific results on the phase 3 clinical trials as well as general knowledge concerning vaccines. The second hypothesis is stronger, and some could read it as modelling an extreme optimistic case. Such assumption is based on some very recent experimental results [43, 71] and experimental review paper [44] on one of the most complex and intriguing topic concerning SARS-CoV-2: the immunological response associated to it. In particular, Iyer and colleagues [43] showed that the igG response has practically no conversion for a long period after the onset of symptoms, namely only 3 individuals over 90 had igG seroconversion. This very limited fraction of seroconversion can be taken into account (through a coefficient σ, see Section 2.3) as some vaccinated individuals get infected because they had seroconversion of their vaccine-induced immune response. Moreover, in their review paper on T cell immunity to COVID-19 [44], Karlsson and colleagues stressed that: ‘Generation of memory T cells can provide lifelong protection against pathogens. Previous studies have demonstrated that SARS-CoV- and MERS-CoV-specific T cells can be detected many years after infection. Likewise, SARS-CoV-2-specific CD4+ and CD8+ T cells are distinguished in a vast majority of convalescent donors (*…*). Preliminary results from the two major mRNA vaccine trials in humans have demonstrated potent Th1 responses.’*

### 2.2 Modelling transmission

Global research on how SARS-CoV-2 is transmitted continues to be conducted at time of writing this paper. It is believed that infected people appear to be most infectious just before (around 1-2 days before) they develop symptoms (i.e. in the post-latency stage) and early in their illness [74]. Recent investigations confirmed that *pre-symptomatic* transmission was more frequent than symptomatic transmission [4]. The possibility of contagion from a truly asymptomatic COVID-19 infected person (i.e. an infected individual who does not develop symptoms) is still a controversial matter. However it has been shown that little to no transmission may occur from truly asymptomatic patients [4].

In our model the routes of transmission from COVID-19 patients are included in the *Force of Infection* (FoI) function, i.e. the per capita rate at which susceptibles contract the infection. As in [36], the mass action incidence is considered:

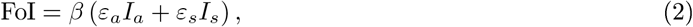

where 0 ≤ *ε*_*a*_, *ε*_*s*_ < 1.

The rationale for this choice is that during observed COVID-19 outbreaks the total population has remained effectively constant. For instance, in Italy (one of the countries more hit by the epidemic [75]), the drop in the total population (≈ 60 · 10^6^) due to the disease-induced deaths (≈ 117 · 10^3^ as of 19 April 2021 [41]) is around 0.195%. In this case, we expect mass action and standard incidence to give similar results.

In (2) the parameters *ε*_*a*_ and *ε*_*s*_ are modification factors that represent the level of reduced infectiousness of compartments *I*_*a*_ and *I*_*s*_ when compared with the subgroup of *I*_*a*_ given by post-latent individuals. Therefore, the baseline transmission rate *β* is the transmission rate of post-latent individuals (see also Section 4.2 where *ε*_*a*_ and *ε*_*s*_ are estimated). For the reasons discussed above we assume that the factor concerning the post-latent individuals is 1.

### 2.3 Description of the balance equations

All the state variables decrease by natural death, with rate *μ*. The susceptible population *S* increases by the net inflow Λ, incorporating both new births and immigration and decreases due to transmission and vaccination. For the time span covered in our simulations, demography could be neglected. However, including a net inflow of susceptible individuals into the model allows one to consider not only new births, but also immigration, which plays an important role during COVID-19 epidemics and can be well estimated in some cases [8]. Therefore, since the demography parameters can be easily obtained from data, we prefer to use an SEIR-like model with demography as successfully done for SARS models [37]. The exposed (or latent) individuals *E* arise as the result of new infections of susceptible and vaccinated individuals and decrease by development at the infectious stage (at rate *ρ*). We assume that after the end of the latency period, the individuals enter in the asymptomatic compartment *I*_*a*_, which includes post-latent and truly asymptomatic, as described in Section 2.1. Asymptomatic individuals *I*_*a*_ diminish because they enter the compartment of symptomatic individuals *I*_*s*_ (at a rate *η*) or they recover (at a rate *ν*_*a*_). Mildly or severely symptomatic individuals *I*_*s*_ come from the post-latency stage and get out due to recovery (at rate *ν*_*s*_) or disease-induced death (at rate *δ*). Vaccinated individuals *V* come from the susceptible class after vaccination (at least one dose of COVID-19 vaccine) and decrease due to infections (at a reduced rate *σβ*, where *σ* ∈ [0, 1)). Finally, recovered individuals come from the infectious compartments *I*_*a*_ and *I*_*s*_ and, as discussed in Remark 1, acquire long lasting immunity against the disease.

### 2.4 The equations

According to the description above, the time evolution of the state variables is ruled by the following system of balance equations:

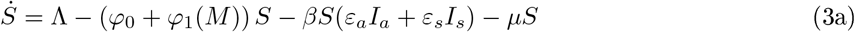

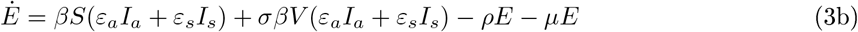

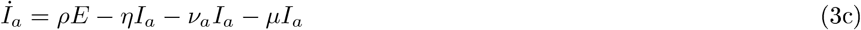

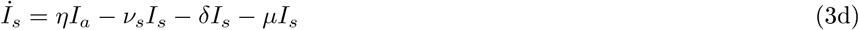

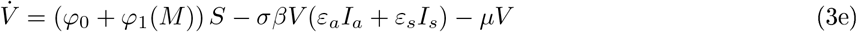

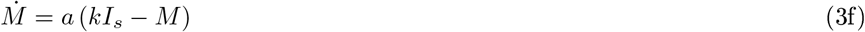

with initial conditions

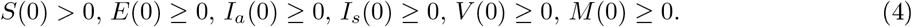

Since the equations (3) do not depend on *R*, the dynamics of the removed compartment can possibly be studied separately, by means of equation

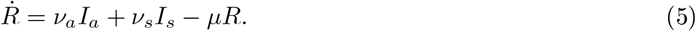

In (3) it is assumed that *φ*_0_ > 0 and *φ*_1_(·) is a continuous increasing function of the information index *M* with *φ*_1_(0) = 0 and sup(*φ*_1_) < 1 − *φ*_0_. The parameter *φ*_0_ embeds: i) the fact that some categories of subjects such as patients and healthcare workers in senior care facilities will be strongly recommended to get the vaccine (and in some countries their vaccination will be even mandatory [48]); ii) the fact that some people are strongly in favour of vaccines and act coherently by getting vaccinated.

The flow chart in Fig. 1 illustrates all the processes included in the model; a description of each parameter together with their baseline values is given in Table 1 (see Section 4).

**Table 1:** Temporal horizon, initial conditions and parameters baseline values for model (3)-(23).

| Parameter | Description | Baseline value |
| --- | --- | --- |
| $t_f$ | Time horizon | 365 – 395 days |
| $N_0$ | Initial total population | $6.036 \cdot 10^7$ |
| $E(0)$ | Initial number of exposed individuals | $I_a(0)(\eta + \nu_a + \mu)/\rho$ |
| $I_a(0)$ | Initial number of asymptomatic infectious individuals | 7,322 |
| $I_s(0)$ | Initial number of symptomatic infectious individuals | 7,545 |
| $V(0)$ | Initial number of vaccinated individuals | 0 |
| $R(0)$ | Initial number of recovered individuals | 203,968 |
| $M(0)$ | Initial value of the information index | $kI_s(0)$ |
| $\mathcal{R}_0$ | Basic reproduction number | 1.428 |
| $\mathcal{R}_V$ | Control reproduction number | 0.302 |
| $\Lambda$ | Net inflow of susceptibles | $1,762 \text{ days}^{-1}$ |
| $\mu$ | Natural death rate | $1.07 \cdot 10^{-2} \text{ years}^{-1}$ |
| $\beta$ | Baseline transmission rate | $2.699 \cdot 10^{-8} \text{ days}^{-1}$ |
| $q$ | Fraction of post-latent individuals that develop symptoms | 0.15 |
| $\varepsilon_a$ | Modification factor concerning transmission from $I_a$ | $q + (1 - q)0.033$ |
| $\varepsilon_s$ | Modification factor concerning transmission from $I_s$ | 0.034 |
| $\varphi_0$ | Information-independent constant vaccination rate | $0.002 \text{ days}^{-1}$ |
| $\sigma$ | Factor of vaccine ineffectiveness | 0.2 |
| $\rho$ | Latency rate | $1/5.25 \text{ days}^{-1}$ |
| $\eta$ | Rate of onset of symptoms | $0.12 \text{ days}^{-1}$ |
| $\nu_a$ | Recovery rate for asymptomatic infectious individuals | $0.165 \text{ days}^{-1}$ |
| $\nu_s$ | Recovery rate for symptomatic infectious individuals | $0.055 \text{ days}^{-1}$ |
| $\delta$ | Disease-induced death rate | $6.248 \cdot 10^{-4} \text{ days}^{-1}$ |
| $D$ | Reactivity factor of information-dependent vaccination | $500\mu/\Lambda$ |
| $\varphi_{max}$ | Ceiling of overall vaccination rate | $0.02 \text{ days}^{-1}$ |
| $a$ | Inverse of the average information delay $T_a$ | $1/3 \text{ days}^{-1}$ |
| $k$ | Information coverage | 0.8 |

**Figure 1:**
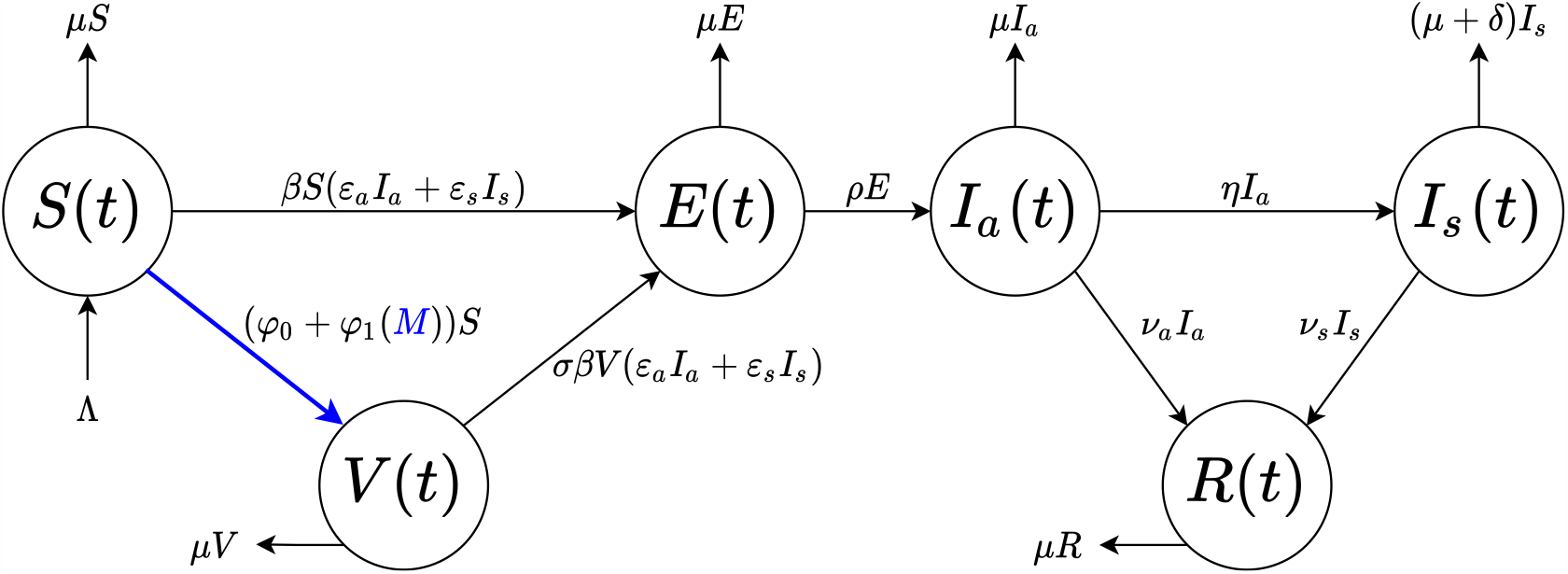
Flow chart for the COVID-19 model (3)-(5). The population *N*(*t*) is divided into six disjoint compartments of individuals: susceptible *S*(*t*), exposed *E*(*t*), asymptomatic *I*_*a*_(*t*), symptomatic *I*_*s*_(*t*), vaccinated *V* (*t*) and recovered *R*(*t*). Blue colour indicates the information-dependent process in the model, with *M* (*t*) ruled by (3f).

## 3 Qualitative analysis

The following theorem ensures that the solutions of model (3) are epidemiologically and mathematically well-posed.

### Theorem 1.

*The region* 𝒟 *defined by*

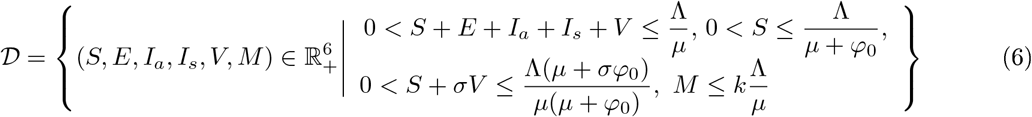

*with initial conditions (4) is positively invariant for model (3)*.

*Proof*. By standard procedure (see e.g. [66]), from (3)-(4) one can derive that

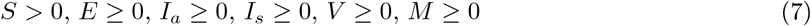

for all *t ≥* 0.

Let us introduce the variable *Ñ* = *S + E +I*_*a*_ *+I*_*s*_ *+V*, that is, at each time *t*, the total population devoid of the removed individuals. Adding the first five equations of the system (3), we obtain

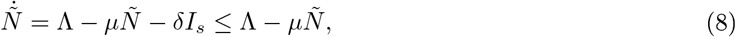

where we use (7). The solution *Ñ* of the differential equation in (8) has the following property

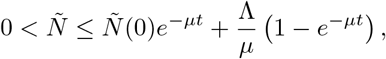

implying that 0 < *Ñ* ≤ Λ*/μ*, as *t →* +∞. Specifically, if *Ñ* (0) *≤*Λ*/μ*, then Λ*/μ* is the upper bound of *Ñ*; if *Ñ* (0) > Λ*/μ*, then *Ñ* will decrease to Λ*/μ*.

Similarly, from equation (3a) and property (7), it follows that 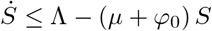, yielding

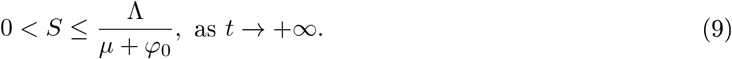

Then,

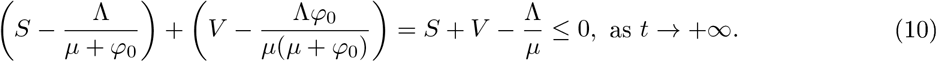

Inequalities (9) and (10), taking into account that *σ ∈* [0, 1), imply that

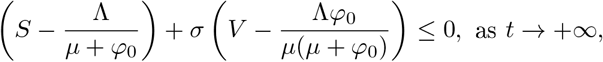

namely 0 < *S* + *σV* ≤ Λ(*μ* + *σφ*_0_)*/*(*μ*(*μ* + *φ*_0_)), as *t →* +∞.

Let us now prove that *M* ≤ *k*Λ*/μ*, as *t →* +∞. From the definition of *M*, as given in (1), it easily follows

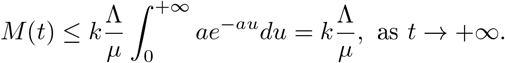

This completes the proof that the region 𝒟, as defined in (6), is positively invariant under the flow induced by the system (3).

Thus, it is not restrictive to limit our analyses to the region *𝒟*.

### 3.1 Disease-free equilibrium and its stability

The model given by equations (3) has a unique disease-free equilibrium (DFE), obtained by setting the r.h.s. of equations (3) to zero, given by

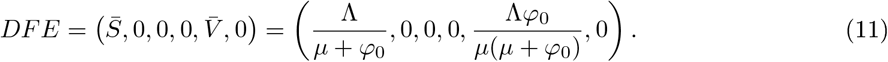

To establish the local and global stability of the DFE, suitable threshold quantities are computed: the basic and control reproduction numbers. The *basic reproduction number*, ℛ_0_, is a frequently used indicator for measuring the potential spread of an infectious disease in a community. It is defined as the average number of secondary cases produced by one primary infection over the course of the infectious period in a fully susceptible population. If the system incorporates vaccination strategies, then the corresponding quantity is named the *control reproduction number* and is usually denoted by ℛ_*V*_.

The reproduction number can be calculated as the spectral radius of the *next generation* matrix FV^−1^, where F and V are defined as Jacobian matrices of the new infection appearance and the other rates of transfer, respectively, calculated for infected compartments at the disease-free equilibrium [70]. In this specific case, if *φ*_0_ + *φ*_1_(*M*) = 0 in (3), namely when a vaccination program is not in place, we obtain the expression of *ℛ*_0_; otherwise, the corresponding *ℛ*_*V*_ can be computed.

#### Theorem 2.

*The basic reproduction number of model (3) is given by*

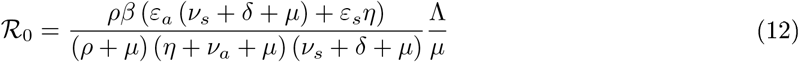

*and the control reproduction number is given by*

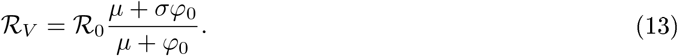

*Proof*. Following the procedure and the notations adopted by Diekmann *et al*. [21] and Van den Driessche & Watmough [70], we derive the control reproduction number, ℛ_*V*_.

Let us consider the r.h.s. of equations (3b)-(3c)-(3d) (the balance equations for the infected compartments), and distinguish the new infections appearance from the other rates of transfer, by defining the vectors

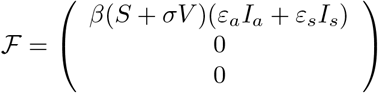

and

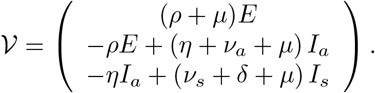

The Jacobian matrices of *ℱ* and *𝒱* evaluated at model DFE (11) read, respectively,

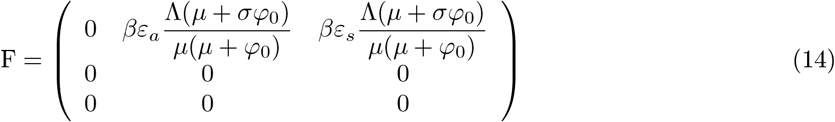

and

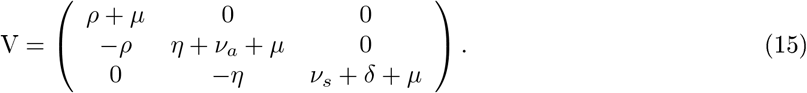

As proved in [21,70], the control reproduction number is given by the spectral radius of the *next generation* matrix FV^−1^. It is easy to check that FV^−1^ has positive elements on the first row, being the other ones null. Thus, *ℛ*_*V*_ = (FV^−1^)_11_, that is

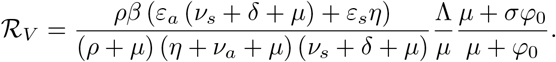

Similarly one can prove that the basic reproduction number is given by (12).

From Theorem 2, it follows that [70]:

#### Proposition 1.

*The DFE is locally asymptotically stable if ℛ*_*V*_ < 1; *otherwise, if ℛ*_*V*_ > 1, *it is unstable*. As far as the global stability of the DFE, we prove the following theorem

#### Theorem 3.

*The DFE is globally asymptotically stable (GAS) if ℛ*_*V*_ < 1.

*Proof*. To prove the global stability of the DFE, we adopt the approach developed by Castillo-Chavez *et al*. in [12]. We rewrite system (3) in the form

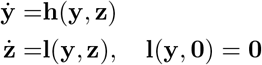

where **y** = (*S, V, M*) denotes the vector of uninfected compartments and **z** = (*E, I*_*a*_, *I*_*s*_) that of infected compartments. The disease-free equilibrium (11) is also rewritten as 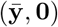, with 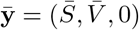 and **0** *∈*ℝ^3^. Then, the DFE is globally asymptotically stable if ℛ_*V*_ < 1, provided that the two following conditions are satisfied [12]:

C.1 For 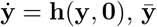 is GAS.

C.2 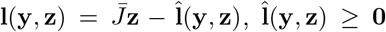 in 𝒟, where 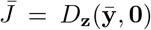 is an M-matrix (the off-diagonal elements are non-negative).

Condition C.1 is immediate, since 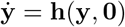 reads

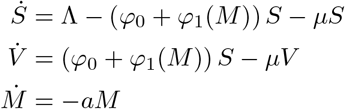

yielding

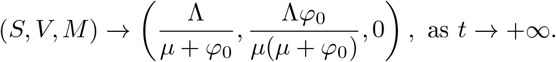

The matrix 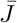 is given by 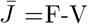, with F and V as computed in the proof of Theorem 2 and given in (14) and (15), respectively. It is easily follows that 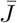 is an M-matrix. Further, in view of (6),

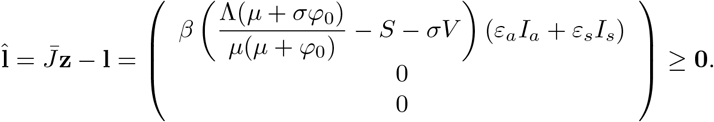

Hence, also condition C.2 is satisfied and the proof is completed.

For an alternative proof see Appendix A.

We remark that by introducing

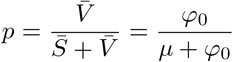

as the fraction of the population vaccinated at the disease-free equilibrium (11) we can express

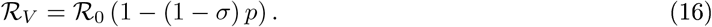

Note that *ℛ*_*V*_ ≤*ℛ*_0_ with equality only if *φ*_0_ = 0 (i.e., *p* = 0) or *σ* = 1. That is, despite being imperfect, the vaccine (characterized by *φ*_0_ > 0 and 0 ≤*σ* < 1) will always reduce the reproduction number of the disease.

The expression (16) is the same as obtained by Gumel *et al*. [36] for the SARS epidemic control. In [36], a detailed analysis is given, leading to the following main results:

#### Proposition 2.

*The disease will be eliminated from the community if p ≥ p*_*c*_, *with p*_*c*_ *given by*

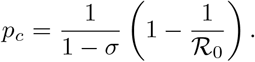

#### Proposition 3.

*Let us consider the following quantity:*

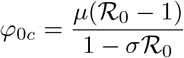

*We have that: if ℛ*_0_ < 1*/σ and φ*_0_ > *φ*_0*c*,_ *then the disease will eliminate from the community. If ℛ*_0_ *≥* 1*/σ, then no amount of vaccination will prevent a disease outbreak in the community*.

See also Fig. 5 in [36], where the critical value, *p*_*c*_, is plotted as a function of 1 − *σ* for several values of *ℛ*_0_.

### 3.2 Endemic equilibrium

Let us denote the generic endemic equilibrium (EE) of model (3) with

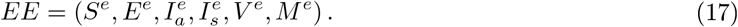

By setting the r.h.s. of equations (3b)-(3c)-(3d)-(3e)-(3f) to zero, one can derive the relationships

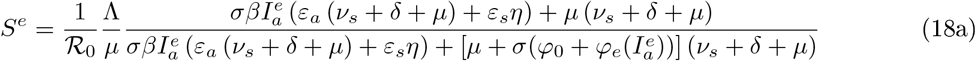

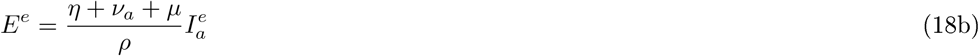

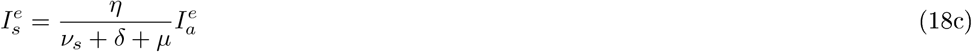

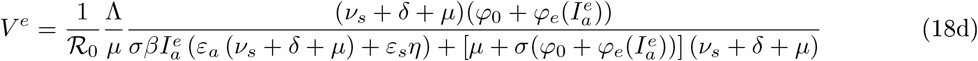

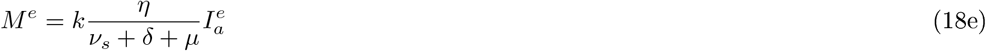

where

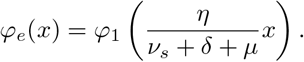

By substituting 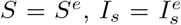 and *M* = *M* ^*e*^ in the r.h.s. of equation (3a) and setting it to zero, we obtain 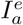 as a positive solution (when it exists) of

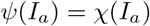

where

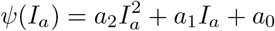

with

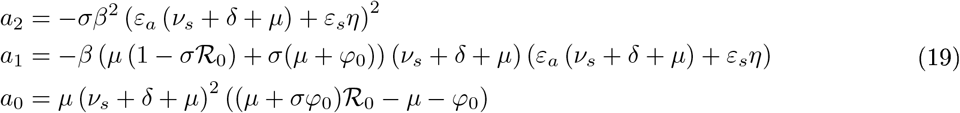

and

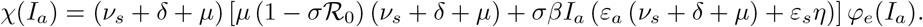

In view of (6), we can limit ourselves to seek 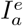 in the interval (0, Λ*/μ*).

Firstly, let us list some proprieties of the functions *ψ*(*I*_*a*_) and *χ*(*I*_*a*_), that can be easily verified:

i. *ψ* (*I*_*a*_) is a concave quadratic function;
ii. *χ*(*I*_*a*_) is the product of a linear-affine increasing function and a positive increasing function (*φ*_*e*_(·));
iii. sgn(*ψ*(0)) = sgn(*ℛ* _*V*_ − 1) and *χ*(0) = 0;
iv. *ψ* (Λ*/μ*) < 0 < *χ*(Λ*/μ*);
v. 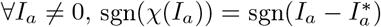, where

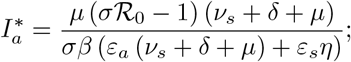
vi. 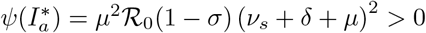.

Then, we distinguish three cases:

- *ℛ* _0_ ≤ (*μ* + *φ*_0_)*/*(*μ* + *σφ*_0_) (namely, *ℛ* _*V*_ ≤ 1). Then,

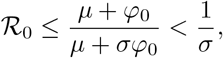

implying that *a*_0_ = *ψ*(0) ≤ 0, *a*_1_ = *ψ′*(0) < 0 and *χ*(*I*_*a*_) is increasing and positive *∀I*_*a*_ > 0. From (i)-(iii) it follows that *ψ*(*I*_*a*_)and *χ*(*I*_*a*_) cannot intersect for *I*_*a*_ > 0, namely no endemic equilibria exist.
- (*μ* + *φ*_0_)*/*(*μ* + *σφ*_0_) < *ℛ* _0_ ≤ 1*/σ*. Then, *a*_0_ = *ψ*(0) > 0, *a*_1_ = *ψ′*(0) < 0 and *χ*(*I*_*a*_) is a positive increasing function ∀*I*_*a*_ > 0. From (i)-(iii)-(iv) it follows that *ψ*(*I*_*a*_) and *χ*(*I*_*a*_) have one positive intersection point and it is in (0, Λ*/μ*), namely an unique endemic equilibrium exists.
- ℛ_0_ > 1*/σ*. Then, *a*_0_ = *ψ*(0) > 0 and *χ*(*I*_*a*_) is negative for 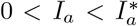 and it is positive and increasing for 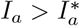. Further,

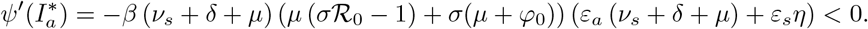

From (i)-(iii)-(iv)-(vi) it follows that *ψ*(*I*_*a*_) and *χ*(*I*_*a*_) have one positive intersection point and it is in 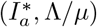, namely an unique endemic equilibrium exists.

Hence, EE exists if and only if *ℛ*_*V*_ > 1 and the endemic number of asymptomatic individuals 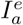 is characterized by 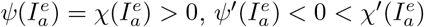 and

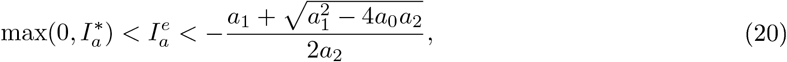

where the last term in (20) is the (unique) positive root of *ψ*(*I*_*a*_).

The results are summarized in the following theorem.

#### Theorem 4.

If *ℛ*_*V*_ ≤ 1, *system (3) admits no endemic equilibria*.

If *ℛ*_*V*_ > 1, *system (3) admits an unique endemic equilibrium, defined in (17)-(18), with* 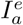 *such that*

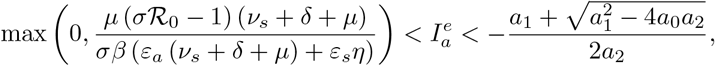

*and a*_*i*_, *i* = 0, …, 2, *given in (19)*.

### 3.3 Central manifold analysis

To derive a sufficient condition for the occurrence of a transcritical bifurcation at ℛ_*V*_ = 1, we can use a bifurcation theory approach. We adopt the approach developed in [27,70], which is based on the general center manifold theory [35]. In short, it establishes that the normal form representing the dynamics of the system on the central manifold is given by:

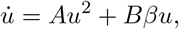

where

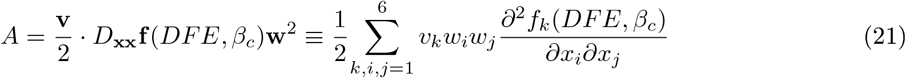

and

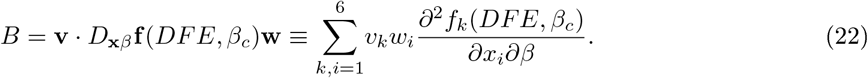

Note that in (21) and (22) *β* has been chosen as bifurcation parameter, *β*_*c*_ is the critical value of *β*, **x** = (*S, E, I*_*a*_, *I*_*s*_, *V,M*) is the state variables vector, **f** is the right-hand side of system (3), and **v** and **w** denote, respectively, the left and right eigenvectors corresponding to the null eigenvalue of the Jacobian matrix evaluated at criticality (i.e. at DFE and *β* = *β*_*c*_).

Observe that *ℛ*_*V*_ = 1 is equivalent to:

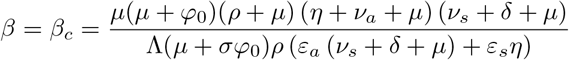

so that the disease-free equilibrium is stable if *β* < *β*_*c*_, and it is unstable when *β* > *β*_*c*_.

The direction of the bifurcation occurring at *β* = *β*_*c*_ can be derived from the sign of coefficients (21) and (22). More precisely, if *A* > 0 [resp. *A* < 0] and *B* > 0, then at *β* = *β*_*c*_ there is a backward [resp. forward] bifurcation.

For our model, we have the following:

#### Theorem 5.

*System (3) exhibits a forward bifurcation at DFE and ℛ*_*V*_ = 1.

*Proof*. The Jacobian of system (3) is

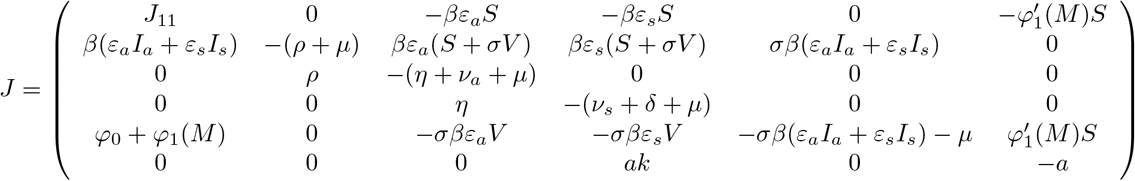

with *J*_11_ = −*β*(*ε*_*a*_*I*_*a*_ + *ε*_*s*_*I*_*s*_) − (*μ* + *φ*_0_ + *φ*_1_(*M*)).

*J* evaluated at DFE (11) for *β* = *β*_*c*_ becomes:

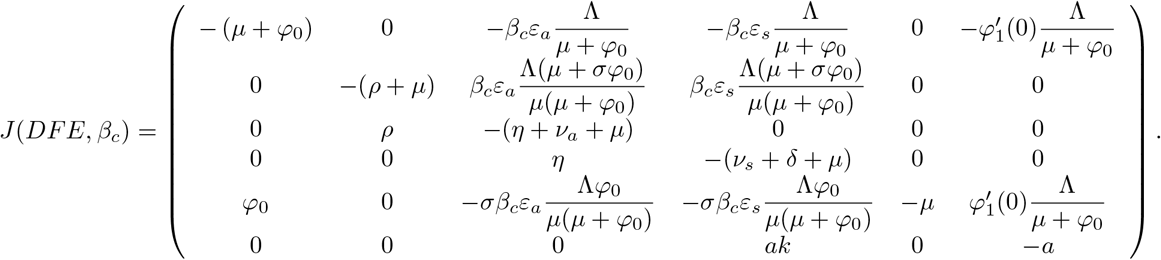

Its spectrum is: Σ = {0, −(*μ* + *φ*_0_), −*μ*, −*a, λ*_+_, *λ*_−_}, where *λ*_*±*_ are given by

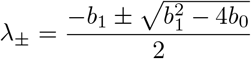

with

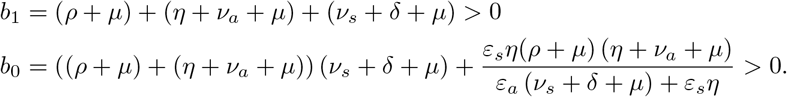

As expected, it admits a simple zero eigenvalue and the other eigenvalues have negative real part. Hence, when *β* = *β*_*c*_ (or, equivalently, when *ℛ*_*V*_ = 1), the DFE is a non-hyperbolic equilibrium.

It can be easily checked that a left and a right eigenvector associated with the zero eigenvalue so that **v**·**w** = 1 are:

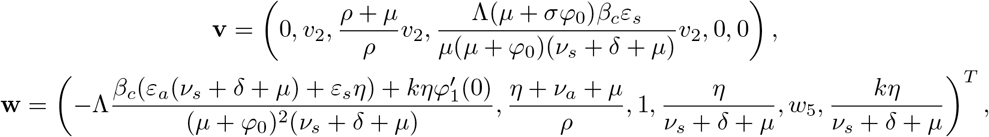

with

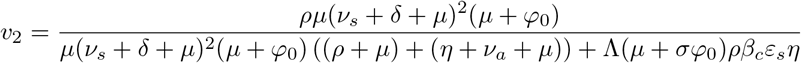

and

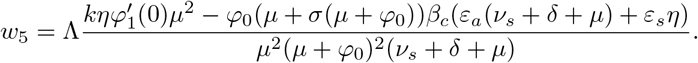

The coefficients *A* and *B* may be now explicitly computed. Considering only the non-zero components of the eigenvectors and computing the corresponding second derivative of **f**, it follows that:

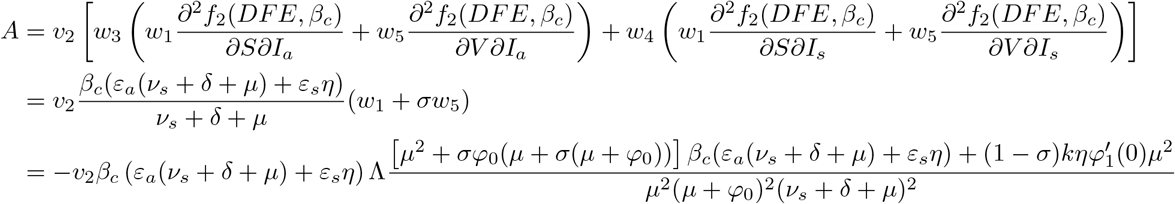

and

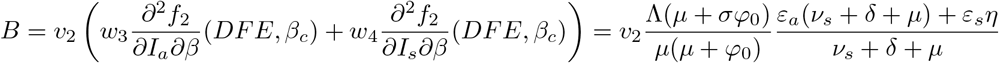

where *v*_2_ > 0. Then, *A* < 0 < *B*. Namely, when *β* − *β*_*c*_ changes from negative to positive, DFE changes its stability from stable to unstable; correspondingly a negative unstable equilibrium becomes positive and locally asymptotically stable. This completes the proof.

## 4 Parametrization

Demographic and epidemiological parameter values are based on the COVID-19 epidemic in Italy reported since the end of February 2020 [41]. Vaccine-related parameter values are mainly inferred by preliminary reports about anti COVID-19 vaccines and by the initial trend of the Italian immunization campaign. A detailed derivation of such quantities is reported in the following.

### 4.1 Initial conditions

In order to provide appropriate initial conditions that mark the beginning of an epidemic wave, we make the following considerations. After the first dramatic epidemic wave (February-May 2020) Italy experiences the so-called ‘living with the virus’ period, characterized by a relatively low level of prevalence and loosening of restrictions. But this breathing space ends towards the second half of August 2020, when the virus regained strength and progressively grew its prevalence, marking the arrival of the second wave. Since data available at the beginning of the second wave are reasonably more accurate than those at the epidemic starting time, we consider them as initial data. More specifically, we take the official national data for infectious (*I*_*a*_ + *I*_*s*_) and recovered (*R*) people at 16 August 2020, that is estimated as the first time after the end of the first wave that the effective reproduction number exceeds the threshold 1 [42]. For that period, the Italian National Institute of Health estimates the fraction of asymptomatic individuals w.r.t. the total case as 49.25% about, namely *I*_*a*_(0) = 0.4925(*I*_*a*_(0) + *I*_*s*_(0)) [39]. As far as the initial values of exposed individuals *E* and the information index *M* are concerned, in the absence of exact data, we infer them by the corresponding expressions at the endemic state, as given in (18). Hence, one yields *E*(0) = *I*_*a*_(0)(*η* + *ν*_*a*_ + *μ*)*/ρ* and *M* (0) = *kI*_*s*_(0). Finally, the initial value of susceptible individuals *S* is obtained by subtracting from the total initial population (say, *N*_0_), as given in [8], namely *S*(0) = *N*_0_ − *E*(0) − *I*_*a*_(0) − *I*_*s*_(0) − *R*(0).

### 4.2 Baseline scenario

In the absence of empirical data about vaccinating attitudes, we follow the approach of [7, 8, 26] and assume that *φ*_1_(*M*) is a Michaelis-Menten function [61]

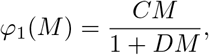

with 0 < *C* ≤ *D*. Similarly to what done in [7, 8, 26], we set *C* = *D* (*φ*_*max*_ − *φ*_0_), where *φ*_*max*_ > *φ*_0_. This reparametrisation means an asymptotic overall rate of *φ*_*max*_ days^−1^. The ensuing vaccination function is:

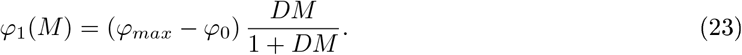

As of April 2021, the rate of anti COVID-19 vaccination in Italy was less than 400,000 administrations *per* day in a population of *N*_0_ ≈ 60 millions of inhabitants [39], but acceleration plans have been laid out. Here, we take *φ*_*max*_ = 0.02 days^−1^ potentially implying a ceiling of 0.02 days^−1^ in vaccination rate under circumstances of high perceived risk. This value is in line with data concerning the 2009 H1N1 pandemic influenza, whose daily rate of vaccine administration has been largely investigated and it was below 2% of the total population (see [49] and references therein). Furthermore, threshold values of 1-2% *per* day were also considered in epidemic models of dengue [67] and cholera diseases [30].

In order to obtain a baseline value for *D*, we observe that in [6, 26] it was set *D* = 500, where *M* varied in [0, *k*]. Here *M* varies in [0, *k*Λ*/μ*] (see (6)), hence we expect that *D* = 500*μ/*Λ could be a good starting point.

As far as the factor of vaccine ineffectiveness, *σ*, and the information-independent constant vaccination rate, *φ*_0_, are concerned, in Section 5 numerical solutions by varying both *σ* ∈ [0, 1) and *φ*_0_ ∈ [0, *φ*_*max*_] are given. Anyway, for illustrative purposes, a corresponding baseline value is selected: *σ* = 0.2, meaning that the vaccine offers 80% protection against infection, and *φ*_0_ = 0.002 days^−1^, that is the 10% of the ceiling vaccination rate *φ*_*max*_ (*φ*_0_ = 0.1*φ*_*max*_). Specifically, 80% is the estimated effectiveness of partial immunization (14 days after first dose but before second dose) of some authorized mRNA COVID-19 vaccines [13].

We estimate the rate at which symptoms onset as *η* = *qγ*, where *q* = 0.15 represents the fraction of infected people that develops symptoms after the incubation period and *γ* = 1*/*1.25 days^−1^ is the post-latency rate, as given in [8]. The fraction *q* is also used to infer *ε*_*a*_, the modification factor concerning transmission from *I*_*a*_, namely we set *ε*_*a*_ = *q* + (1 − *q*)0.033, where 1 [resp. 0.033] is the modification factor concerning transmission from post-latent [resp. truly asymptomatic] individuals, as considered in the models [8, 33].

Following the approach adopted by Gumel *et al*. [37], based on the formula given by Day [17], we estimate the disease-induced death rate as

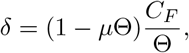

where *C*_*F*_ is the fatality rate and Θ is the expected time from the onset of symptoms until death. We compute *C*_*F*_ by the official national data from 16 August to 13 October 2020 [41] (the same period considered for the estimation of the transmission rate *β*, as explained below), yielding *C*_*F*_ = 0.75%. As far as Θ is concerned, from [39] we get Θ = 12 days, providing *δ* ≈ 6.248 ·10^−4^ days^−1^.

Similarly, the recovery rates *ν*_*j*_ with *j* ∈ {*a, s*} are estimated as

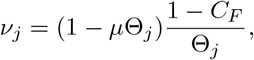

where Θ_*a*_ [resp. Θ_*s*_] is the expected time until recovery for asymptomatic [resp. symptomatic] individuals. We assume Θ_*a*_ = 6, Θ_*s*_ = 18 days on the basis of the considerations made in [8].

Values for Λ, *μ, ε*_*s*_, *ρ, a* and *k* are based on the estimates given in [8]. Like as for *σ* and *φ*_0_, numerical solutions by varying both *k* ∈ [0.2, 1] and *a* ∈ [1*/*60, 1] days^−1^ are given in Section 5 (for a detailed motivation about the ranges of values of the information parameters see [8]).

Finally, in order to obtain an appropriate value for the baseline transmission rate *β*, we consider model (3)-(23) in absence of vaccination strategies (*φ*_0_ = 0 days^−1^, *D* = 0) and search for the value that best fits with the initial ‘uncontrolled’ phase of the second Italian epidemic wave. More precisely, we consider the number of COVID-19-induced deaths in Italy from 16 August, assumed as the starting date of the second wave (see Section 4.1), and 13 October 2020, the last day of loose restrictions. Indeed, on 13 October the Council of Ministers approved a decree to reintroduce stricter rules to limit the spread of the disease [40]. The choice of the curve to fit is motivated by the fact that data about deaths seem to be more accurate with respect to other ones, e.g. the number of infected people, who are not always identified, especially if asymptomatic or with very mild symptoms. Anyway, by setting *β* = 2.699 · 10^−8^ days^−1^, we obtain a good fit not only with the cumulative deaths (see Fig. 2B) but also with the total infectious cases, *I*_*a*_ + *I*_*s*_ (see Fig. 2A).

**Figure 2:**
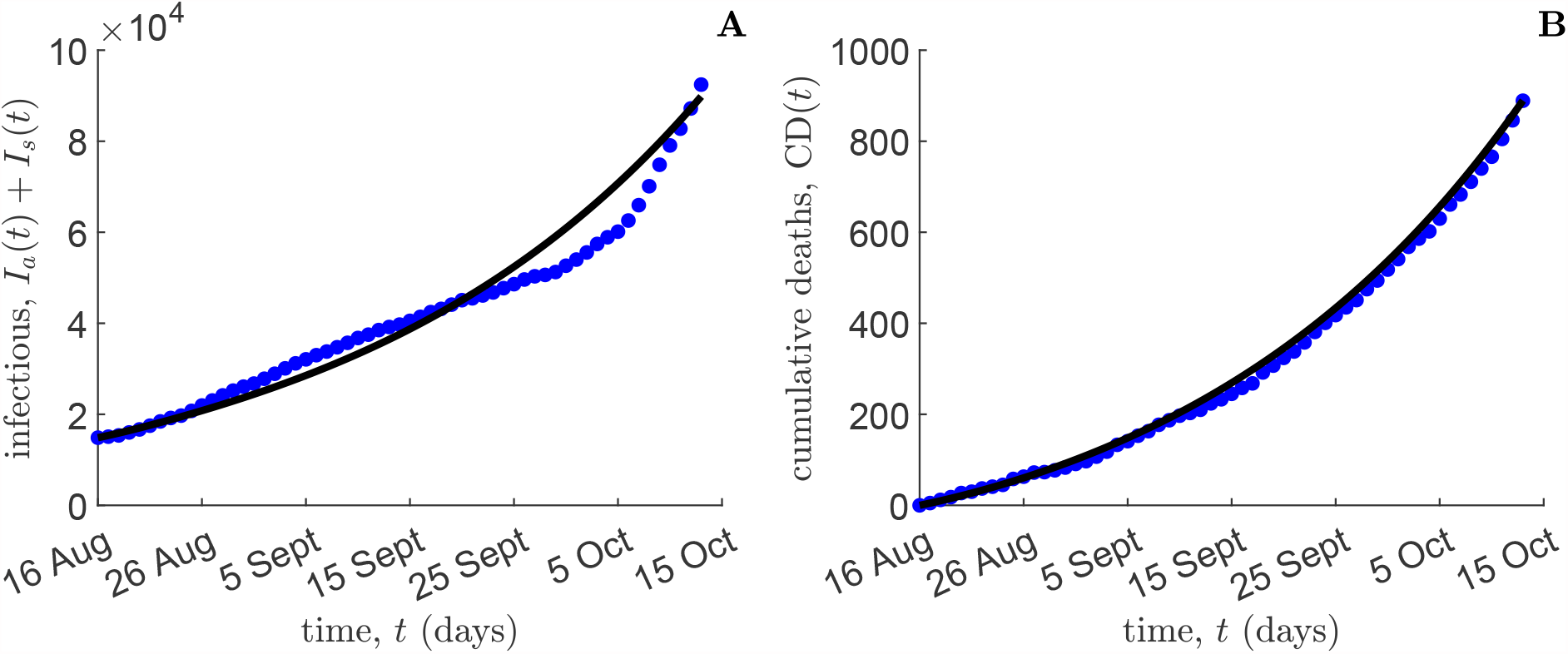
Dynamics in absence of vaccination (*φ*_0_ = 0 days^−1^, *D* = 0). Total infectious cases (panel A) and cumulative disease-induced deaths (panel B) as predicted by model (3)-(23) (black lines) and compared with Italian official data [41] (blue dots), in the period 16 August-13 October 2020. Initial conditions and other parameter values are given in Table 1.

All the parameters of the model as well as their baseline values are reported in Table 1.

## 5 Numerical simulations

Numerical simulations are performed in MATLAB [57]. We use the 4th order Runge-Kutta method with constant step size for integrating the system and the platform-integrated functions for getting the plots. First, we numerically investigate the impact of two vaccine-related parameters, namely the information-independent constant vaccination rate, *φ*_0_, and the factor of vaccine ineffectiveness, *σ*, on the control reproduction number *ℛ*_*V*_ of formula (13). The corresponding contour plot of *ℛ*_*V*_ (*φ*_0_, *σ*) is shown in Fig. 3A. This figure shows that: i) for very small values of *φ*_0_ this parameter impacts on *ℛ*_*V*_ but *φ*_0_ > 0.002 days^−1^ about yields that *ℛ*_*V*_ depends practically only on *σ* in a linear-affine manner as shown in Fig. 3B; ii) for small values of *σ* (as those declared for some of the vaccines) the *ℛ*_*V*_ is small, for example for *σ* = 0.05 it is *ℛ*_*V*_ < 0.1; iii) for values of *σ* ≈ 1*/*3, comparable with those observed often for vaccine against the seasonal flu, it is *ℛ*_*V*_ ≈ 0.5; iv) if we define as threshold of non-effectiveness the curve *ℛ*_*V*_ = 1 we observe that for *φ*_0_ > 0.002 days^−1^ this threshold is reached for values of *σ* between around 0.6 and 0.7.

**Figure 3:**
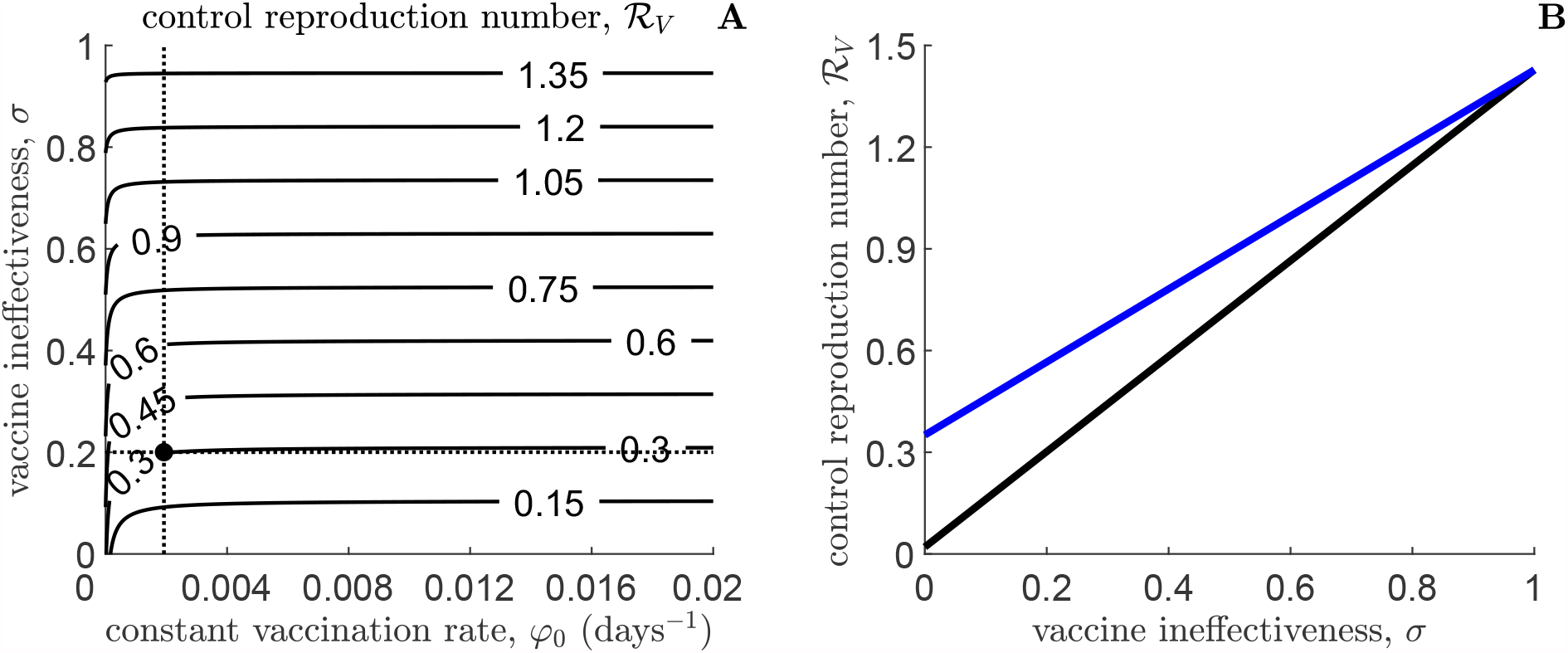
Panel A: Contour plot of the control reproduction number ℛ_*V*_ (13) versus the information-independent constant vaccination rate, *φ*_0_, and the factor of vaccine ineffectiveness, *σ*. Intersection between dotted black lines indicates the value corresponding to the baseline scenario: *φ*_0_ = 0.002 days^−1^, *σ* = 0.2. Panel B: plot of ℛ_*V*_ versus *σ*, by setting *φ*_0_ = 0.002 days^−1^ (black line) and *φ*_0_ = 2·10^−5^ days^−1^ (blue line). Other parameters values are given in Table 1.

### 5.1 Temporal dynamics

Let us consider the time frame [0, *t*], where 0 ≤ *t* ≤*t*_*f*_. We introduce four relevant cumulative quantities that will be used in the following: the cumulative vaccinated individuals CV(*t*), i.e. the total number of individuals who are vaccinated with at least one dose of COVID-19 vaccine in [0, *t*]; the cumulative symptomatic cases CY(*t*), i.e. the number of new cases showing symptoms in [0, *t*]; the cumulative incidence CI(*t*), i.e. the total number of new cases in [0, *t*]; and the cumulative deaths CD(*t*), i.e. the disease-induced deaths in [0, *t*]. For model (3)-(23) we have,respectively:

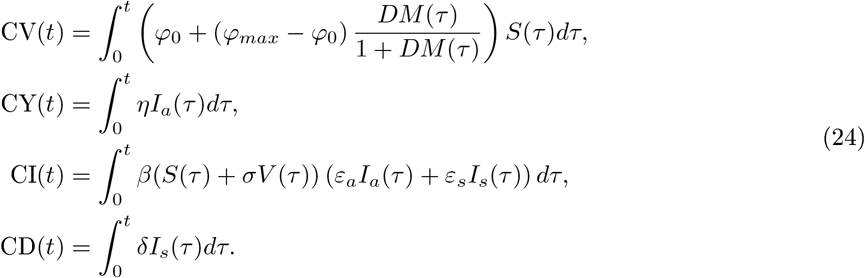

We also consider two possibilities for the time at which vaccines administration starts, namely

- VAX-0, that is the baseline case that the vaccination campaign starts at day *t* = 0;
- VAX-30, that is the case that the vaccination campaign starts at day *t* = 30.

We assume that in both cases the vaccination campaign lasts 1 year, namely *t*_*f*_ = 365 (resp. *t*_*f*_ = 395] days in the case VAX-0 [resp. VAX-30].

Numerical simulations for the case VAX-0 are displayed in Fig. 4. Namely, we report the temporal dynamics of three relevant state variables: susceptible individuals *S* (Fig. 4A), vaccinated individuals *V* (Fig. 4B) and symptomatic infectious individuals *I*_*s*_ (Fig. 4C), as well as the cumulative number of deaths CD (Fig. 4D). We consider the following four significant scenarios (for each of them we also report the observed results):

**Figure 4:**
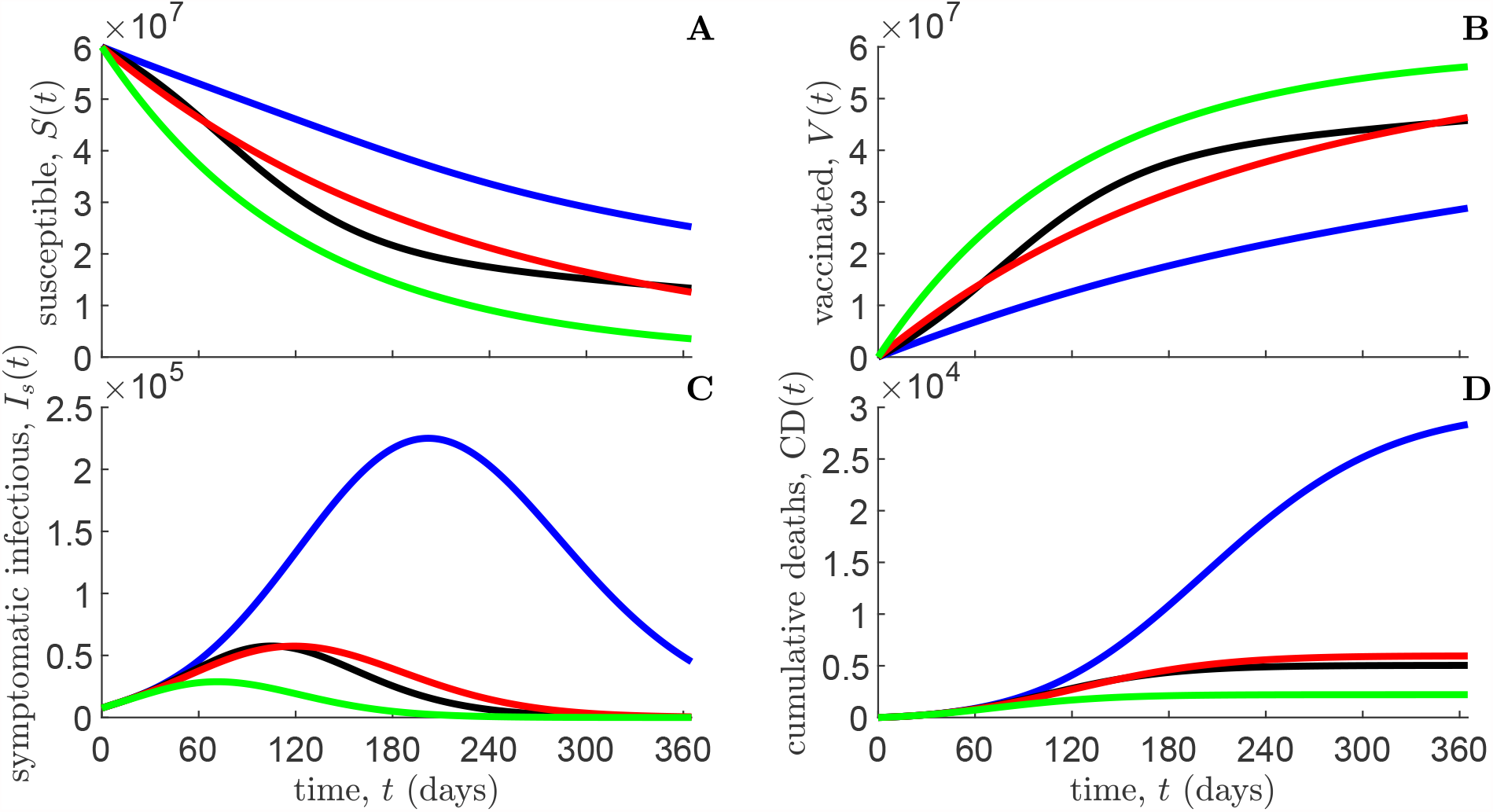
VAX-0 case. Temporal dynamics of susceptible individuals *S* (panel A), vaccinated individuals *V* (panel B), symptomatic infectious individuals *I*_*s*_ (panel C), and cumulative deaths CD (panel D), as predicted by model (3)-(23). Blue lines: constant vaccination with *φ*_0_ = 0.002 days^−1^, *D* = 0; black lines: information-dependent vaccination with *φ*_0_ = 0.002 days^−1^, *D* = 500*μ/*Λ; red lines: constant vaccination with 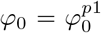, *D* = 0; green lines: constant vaccination with 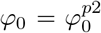, *D* = 0. Initial conditions and other parameter values are given in Table 1 and in Section 5.1.

- Constant vaccination (*D* = 0), with baseline rate *φ*_0_ = 0.002 days^−1^ (blue lines). We observe at *t* = 202 days the occurrence of a large peak of symptomatic cases *I*_*s*_ (225,025) and at the end of simulation a large cumulative number of deaths (28,343);
- Information-dependent vaccination: *φ*_0_ = 0.002 days^−1^, *D* = 500*μ/*Λ (black lines). This case is characterized by a time of *I*_*s*_ peak that is halved w.r.t. the constant baseline case, namely at *t* = 105 days about, and a much lower prevalence: 57, 588, i.e. one quarter about w.r.t. the constant baseline case. This could be an excellent performance, but it is not the case since better performance could have been reached appropriately higher vaccination rate levels;
- Constant vaccination (*D* = 0), with rate 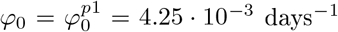 (red lines), which is such that the peak value of *I*_*s*_ is equal to the peak value observed in the case of information-dependent vaccination. One can observe that in this case the epidemic peak occurs earlier, at *t* = 119 days, and the final cumulative number of death is smaller: CD(*t*_*f*_) = 5, 948;
- Constant vaccination (*D* = 0), with rate 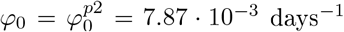 (green lines), where the peak of *I*_*s*_ is halved w.r.t. the case of information-dependent vaccination. The epidemic peak occurs very early, at *t* = 72 days, and the final cumulative number of death is relatively modest: CD(*t*_*f*_) = 2, 203.

Simulations for the case VAX-30 are, of course, graphically similar to those in Fig. 4, hence corresponding plots are here omitted. From a quantitative point of view, in order to compare the results in the case VAX-30 w.r.t. the case VAX-0, we focus on the scenario of information-dependent vaccination and report in Table 2 the value of the following epidemiological indicators (not necessarily in this order): the number of susceptible and vaccinated individuals, and the cumulative quantities (24) at the end of the time horizon *t*_*f*_, the peak of symptomatic cases and its occurrence time. Comparison between the cases VAX-0 and VAX-30 is given though the difference operator:

**Table 2:** Information-dependent vaccination case (*φ*_0_ = 0.002 days^−1^, *D* = 500*μ/*Λ). Relevant quantities as predicted by model (3)-(23) in the case that the vaccination campaign starts at day 0, VAX-0 (first column) and in the case that it starts at day 30, VAX-30 (second column). The third column reports the differences between the values corresponding to the VAX-30 case w.r.t. the case VAX-0. Initial conditions and other parameter values are given in Table 1.

| $X$ | $X\vert _{\text{VAX-0}}$ | $X\vert _{\text{VAX-30}}$ | $X\vert _{\text{VAX-30}} - X\vert _{\text{VAX-0}}$ |
| --- | --- | --- | --- |
| $S(t_f)$ | $1.33 \cdot 10^7$ | $1.09 \cdot 10^7$ | $-2.39 \cdot 10^6$ |
| $V(t_f)$ | $4.58 \cdot 10^7$ | $4.77 \cdot 10^7$ | $1.92 \cdot 10^6$ |
| $CV(t_f)$ | $4.62 \cdot 10^7$ | $4.82 \cdot 10^7$ | $2.00 \cdot 10^6$ |
| $\max(I_s)$ | $5.76 \cdot 10^4$ | $9.14 \cdot 10^4$ | $3.38 \cdot 10^4$ |
| $\arg \max(I_s)$ | 105.14 | 110.61 | 5.47 |
| $CY(t_f)$ | $4.42 \cdot 10^5$ | $6.44 \cdot 10^5$ | $2.02 \cdot 10^5$ |
| $CI(t_f)$ | $1.03 \cdot 10^6$ | $1.51 \cdot 10^6$ | $4.80 \cdot 10^5$ |
| $CD(t_f)$ | $5.04 \cdot 10^3$ | $7.30 \cdot 10^3$ | $2.26 \cdot 10^3$ |

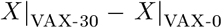

where *X ∈ {S*(*t*_*f*_), *V* (*t*_*f*_), CV(*t*_*f*_), max(*I*_*s*_), arg max(*I*_*s*_), CY(*t*_*f*_), CI(*t*_*f*_), CD(*t*_*f*_) *}* (see third column in Table 2).

Observe that, in both VAX-0 and VAX-30 case, cumulative asymptomatic people at the final time *t*_*f*_ (that is the difference CI(*t*_*f*_)−CY(*t*_*f*_)) account for approximately 57% of cumulative SARS-CoV-2 infections. This result is in line with the current estimates (as of April 2021) reported by the Italian National Institute of Health [39].

We also investigate the temporal dynamics of the ratio *φ*_1_(*M*)*/φ*_0_ in the case of information-dependent vaccination. Numerical solutions are displayed in Fig. 5 for both the case VAX-0 (black line) and the case VAX-30 (blue line). We note that in the case VAX-30 the ratio is larger than in the case VAX-0 since the delay in the start of the vaccination campaign induces a larger epidemic peak. Namely, in the case VAX-0, the maximum value reached by *φ*_1_(*M*)*/φ*_0_ is 2.49 and the time at it is reached is approximately *t* = 108 days. In the case of VAX-30 this peak is reached at *t* = 114, i.e. 84 days after the start of VAX-30, but the peak value is much larger: it is 3.4.

**Figure 5:**
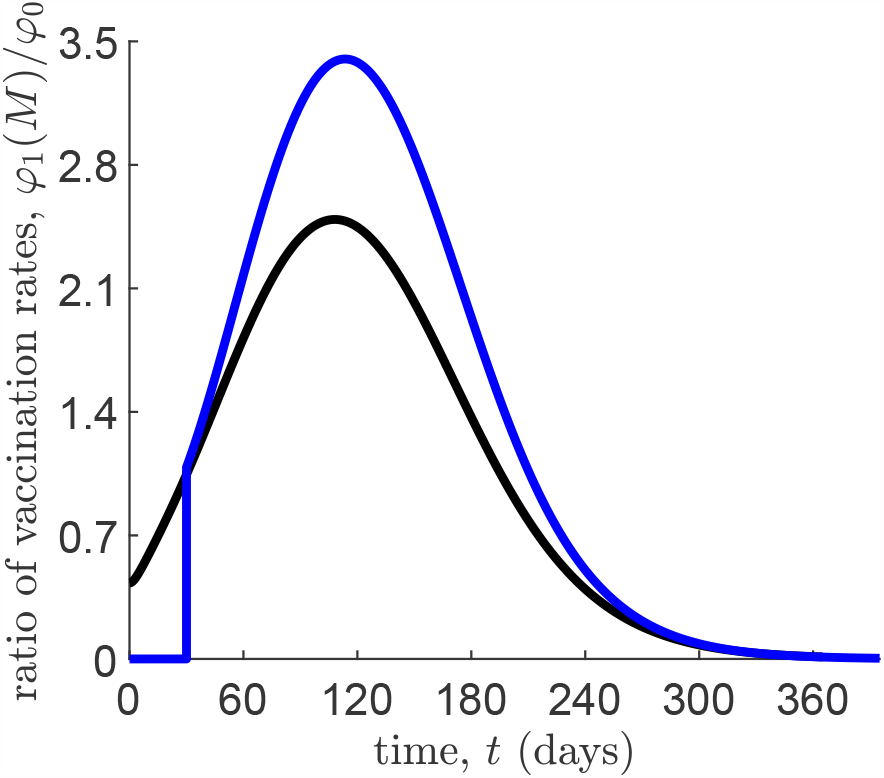
Information–dependent vaccination case (*φ*_0_ = 0.002 days^−1^, *D* = 500*μ/*Λ). Temporal dynamics of the ratio between the information–dependent component, *φ*_1_(*M*), and the constant component, *φ*_0_, of the vaccination rate. Black line: VAX-0 case; blue line: VAX-30 case. Initial conditions and other parameter values are given in Table 1.

### 5.2 Sensitivity of epidemiological indicators to critical parameters

Here, we focus on the VAX-0 case and evaluate the sensitivity of some relevant epidemiological indicators to variations of critical parameter values. Note that for the case VAX-30 we obtain similar results, which we omit.

Specifically, we assess how changing suitable information and vaccine–related parameters affects the cumulative quantities (24) evaluated at the final time *t*_*f*_, the peak of symptomatic cases and its occurrence time. We anticipate here that the final cumulative incidence, CI(*t*_*f*_), the final cumulative symptomatic cases, CY(*t*_*f*_) and the peak of symptomatic cases, max(*I*_*s*_), have in all cases contour plots qualitatively similar to the final cumulative deaths C (*t*_*f*_), thus we do not plot them. Hence, the following figures display the counter plots of just three quantities:

- the cumulative vaccinated individuals at *t*_*f*_ = 365 days, CV(*t*_*f*_);
- the occurrence time of the symptomatic prevalence peak, argmax(*I*_*s*_);
- the cumulative disease-induced deaths at *t*_*f*_ = 365 days, C (*t*_*f*_).

We start by investigating how the information parameters, namely the information coverage, *k*, and the information delay, *T*_*a*_ = *a*^−1^, may affect the epidemic course, see Fig. 6. We observe that for argmax(*I*_*s*_) and C (*t*_*f*_) (as well as max(*I*_*s*_), CI(*t*_*f*_) and CY(*t*_*f*_)) the patterns of the contour plots are similar, and in particular: for small *k* = 0.2 the range of the simulated variable when *T*_*a*_ increases is large, whereas for *k* = 1 the range is restricted and low. The inverse phenomenon is observed for CV(*t*_*f*_): the range is restricted and small for low *k* = 0.2 whereas it is larger for *k* = 1.

**Figure 6:**
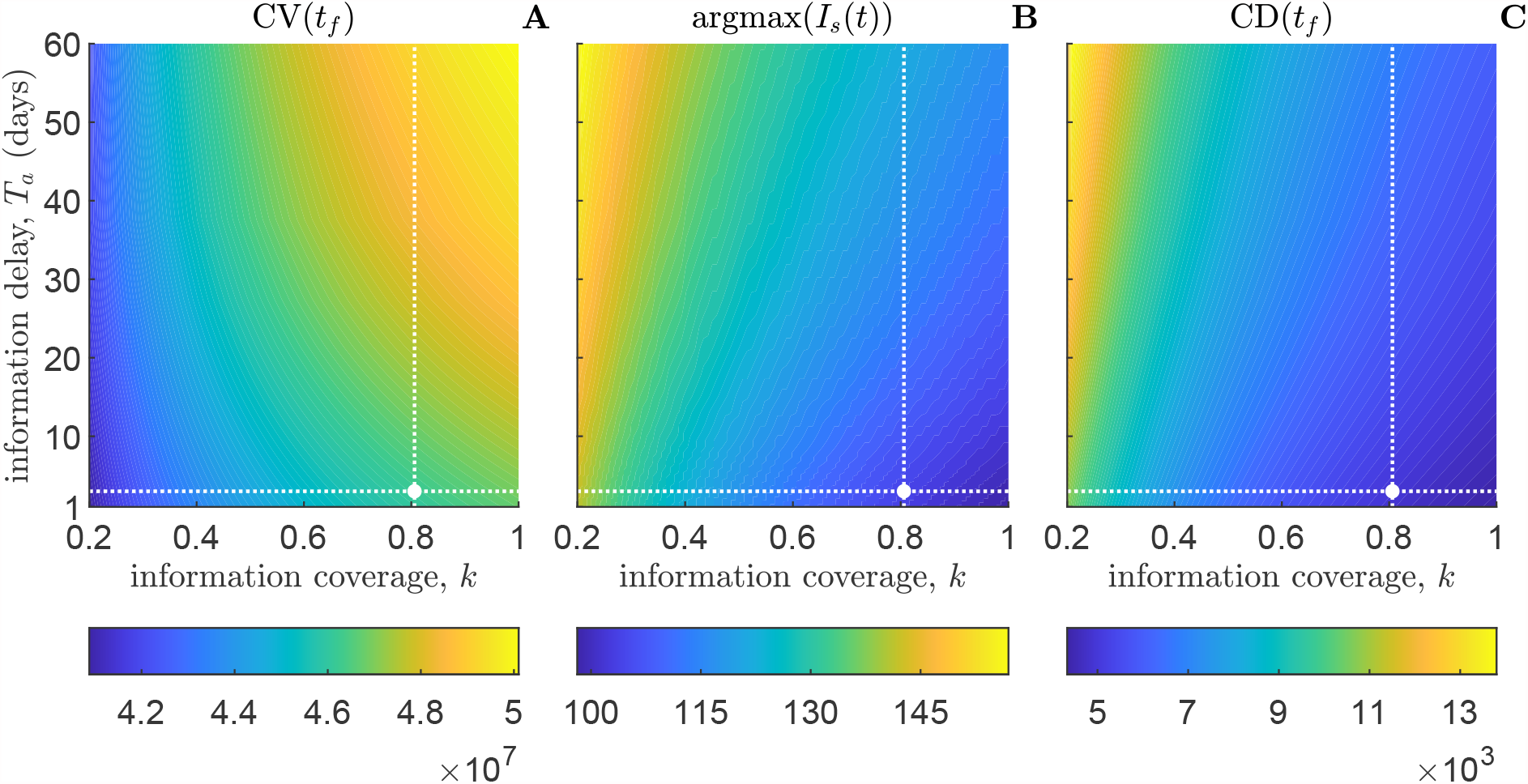
Impact of the information coverage, *k*, and of the average delay, *T*_*a*_ = *a*^−1^, on the VAX-0 scenario as shown by contour plots. Panel A: cumulative vaccinated individuals at the final time *t*_*f*_ = 365 days, CV(*t*_*f*_). Panel B: time of symptomatic prevalence peak, argmax(*I*_*s*_). Panel C: cumulative deaths at the final time *t*_*f*_ = 365 days, CD(*t*_*f*_). The intersection between dotted white lines indicates the values corresponding to the baseline scenario: *k* = 0.8, *T*_*a*_ = 3 days. Initial conditions and other parameter values are given in Table 1.

Then, we investigate how the factor of vaccine ineffectiveness, *σ*, and the information-independent constant vaccination rate, *φ*_0_, affect the same quantities considered above. The results are shown in the contour plots in Fig. 7 for the case of constant baseline vaccination (*φ*_0_ = 0.002 days^−1^, *D* = 0) and in Fig. 8 for the case of information-dependent vaccination (*φ*_0_ = 0.002 days^−1^, *D* = 500*μ/*Λ). We may observe that the quantitative impact of the information-dependent vaccination is remarkable (but this was expected). As far as the shapes of the plots, we note that the plots for CV(*t*_*f*_) (panels A) and for the time at symptomatic prevalence peaks (panels B) are remarkably different from the other plots. Moreover the plot for CV(*t*_*f*_) is qualitatively different in the information–dependent vaccination case w.r.t. the case of constant vaccination.

**Figure 7:**
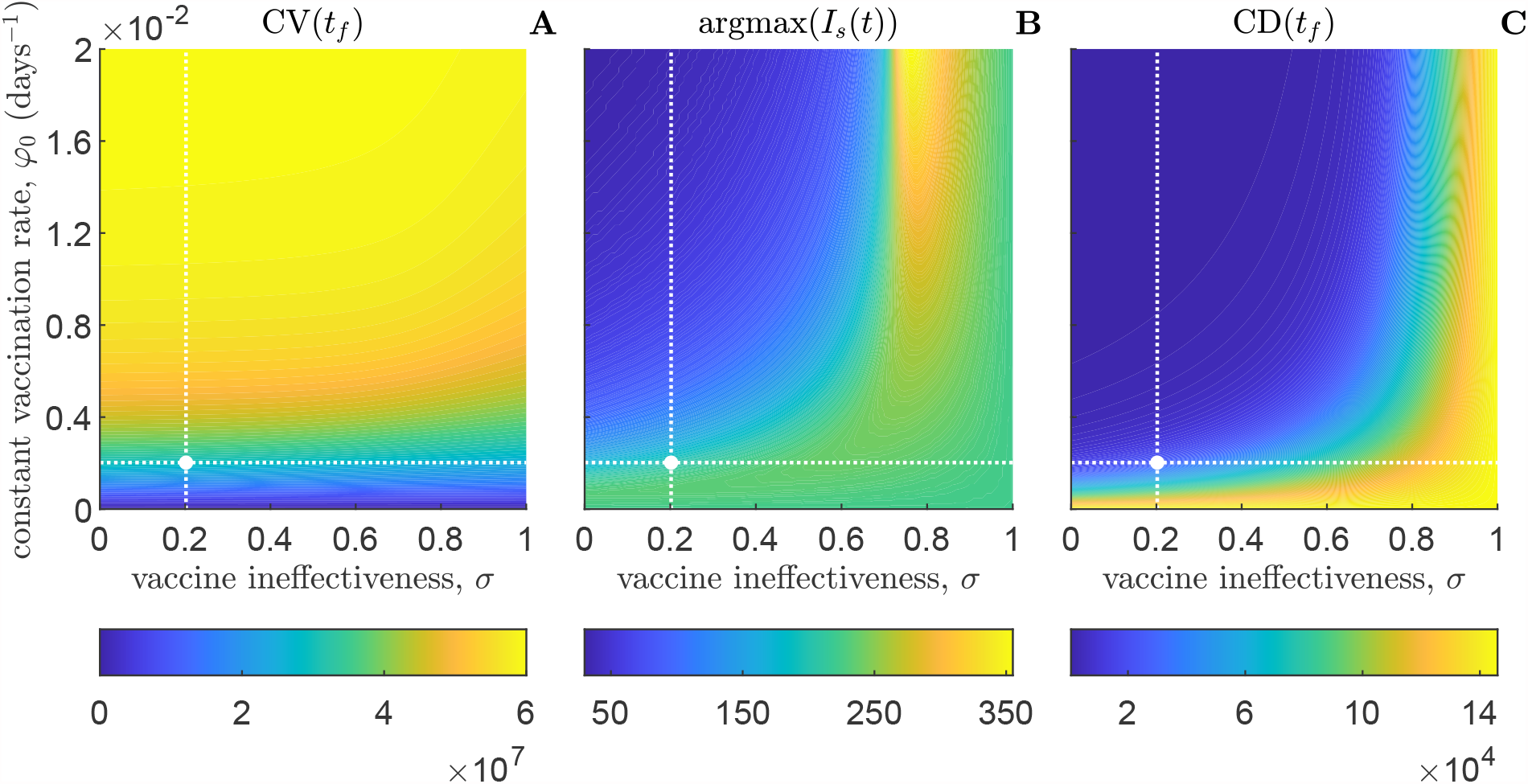
Impact of the factor of vaccine ineffectiveness, *σ*, and of the information–independent constant vaccination rate, *φ*_0_, on the scenario VAX-0 with constant vaccination (i.e. *D* = 500*μ*/Λ) as shown by contour plots. Panel A: cumulative vaccinated individuals at the final time *t*_*f*_ = 365 days, CV(*t*_*f*_). Panel B: time of symptomatic prevalence peak, argmax(*I*_*s*_). Panel C: cumulative deaths at the final time *t*_*f*_ = 365 days, CD(*t*_*f*_). The intersection between dotted white lines indicates the values corresponding to the baseline scenario: *σ* = 0.2, *φ*_0_ = 0.002 days^−1^. Initial conditions and other parameter values are given in Table 1.

**Figure 8:**
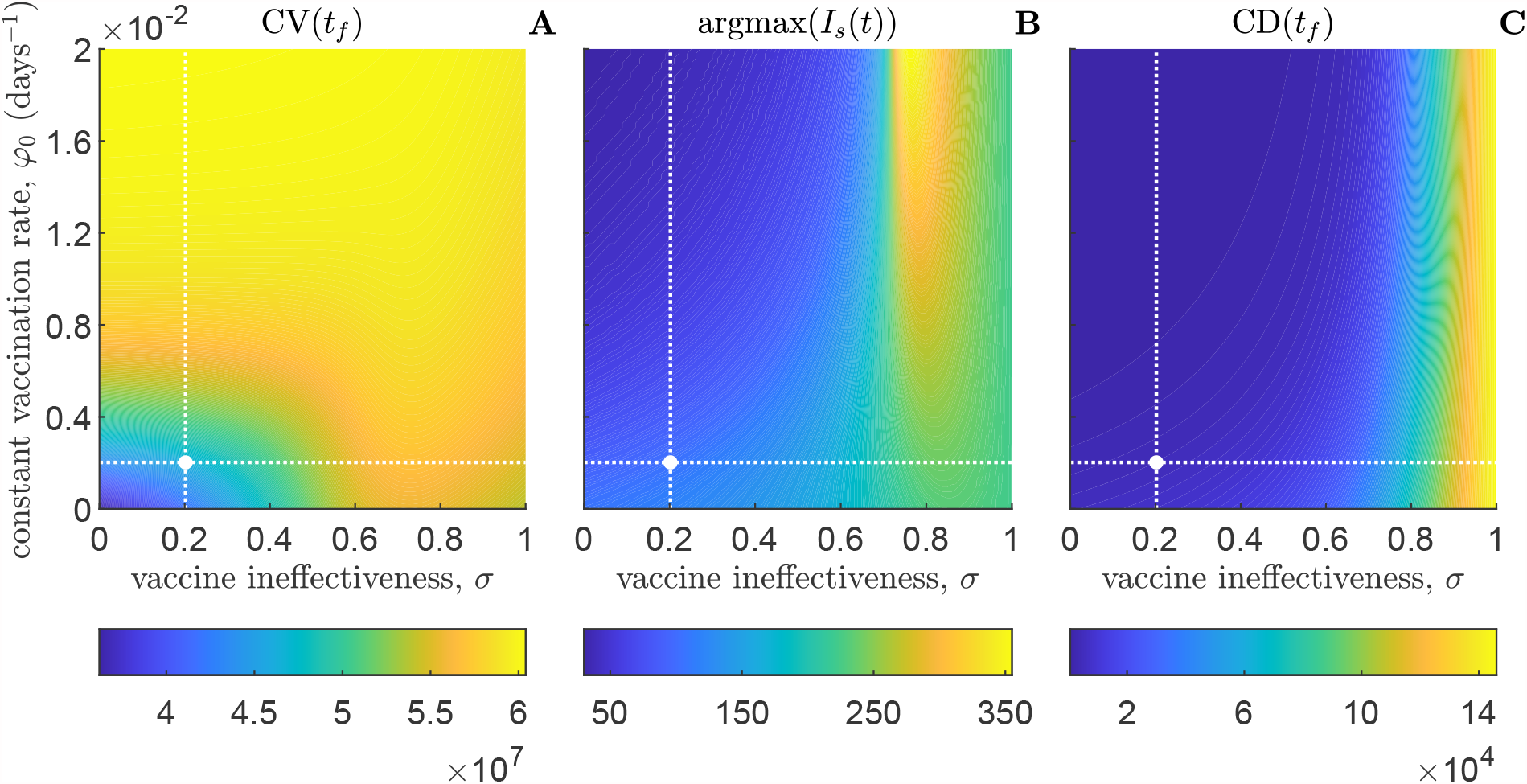
Impact of the factor of vaccine ineffectiveness, *σ*, and of the information–independent constant vaccination rate, *φ*_0_, on the scenario VAX-0 with information-dependent vaccination (i.e. *D* = 500*μ/*Λ), as shown by contour plots. Panel A: cumulative vaccinated individuals at the final time *t*_*f*_ = 365 days, CV(*t*_*f*_). Panel B: time of symptomatic prevalence peak, argmax(*I*_*s*_). Panel C: cumulative deaths at the final time *t*_*f*_ = 365 days, CD(*t*_*f*_). The intersection between dotted white lines indicates the values corresponding to the baseline scenario: *σ* = 0.2, *φ*_0_ = 0.002 days^−1^. Initial conditions and other parameter values are given in Table 1.

## 6 The impact of seasonality

There is an ongoing debate on possible seasonality effects on the transmission and global burden of COVID-19 [1,50,59,65]. Thus, for the sake of the completeness, we consider here the case of information-dependent vaccination and simulate the presence of seasonality on three key parameters: not only the transmission rate, *β*, but also the rate of symptoms onset, *η*, and the total rate of vaccination, *φ*(*M*) = *φ*_0_ + *φ*_1_(*M*), with *φ*_1_(*M*) given in (23). For the latter, the seasonality could be determined by a lower vaccination rate due to the summer vacations.

Namely, we use in our simulations

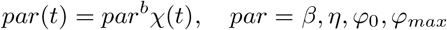

where: *par*^*b*^ are the baseline values and *χ*(*t*) is simply two states switch, i.e. similar to the one proposed in [28] for the transmission rate:

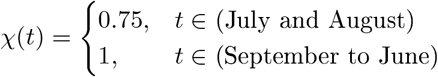

Since we used initial conditions corresponding to COVID–19 data at 16 August 2020, as officially communicated by Italian health authorities (see Section 4.1), we consider:

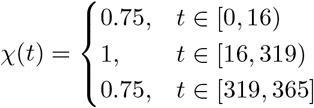

We will denote this simulation scenario with VAX-0S.

Numerical simulations are displayed in Fig. 9 and compared with the baseline scenario, VAX-0. Corresponding relevant quantities are reported in Table 3. Our simulation suggests that: i) the impact of the summer vacation on the vaccine delivery and on *S*(*t*) is minimal (and they are omitted from Fig. 9); ii) the peak of symptomatic cases decreases many months after the summer decrease of the transmission and symptoms onset w.r.t. the no seasonality scenario, and it is delayed (Fig. 9A); iii) the cumulative number of deaths decreases a little bit (Fig. 9B).

**Table 3:** Information–dependent vaccination case (*φ*_0_ = 0.002 days^−1^, *D* = 500*μ*/Λ). Relevant quantities as predicted by model (3)-(23) in the scenario including seasonality VAX-0S (first column). The second column reports the differences between the values corresponding to the VAX-0S case w.r.t. the case VAX-0 (see also Table 2). Initial conditions and other parameter values are given in Table 1 and in Section 6.

| $X$ | $X\vert _{\text{VAX-0S}}$ | $X\vert _{\text{VAX-0S}} - X\vert _{\text{VAX-0}}$ |
| --- | --- | --- |
| $S(t_f)$ | $1.45 \cdot 10^7$ | $1.18 \cdot 10^6$ |
| $V(t_f)$ | $4.47 \cdot 10^7$ | $-1.09 \cdot 10^6$ |
| $CV(t_f)$ | $4.51 \cdot 10^7$ | $-1.12 \cdot 10^6$ |
| $\max(I_s)$ | $5.03 \cdot 10^4$ | $-7.34 \cdot 10^3$ |
| $\arg \max(I_s)$ | 115.66 | 10.52 |
| $CY(t_f)$ | $4.02 \cdot 10^5$ | $-3.97 \cdot 10^4$ |
| $CI(t_f)$ | $9.44 \cdot 10^5$ | $-8.91 \cdot 10^4$ |
| $CD(t_f)$ | $4.59 \cdot 10^3$ | -444.88 |

**Figure 9:**
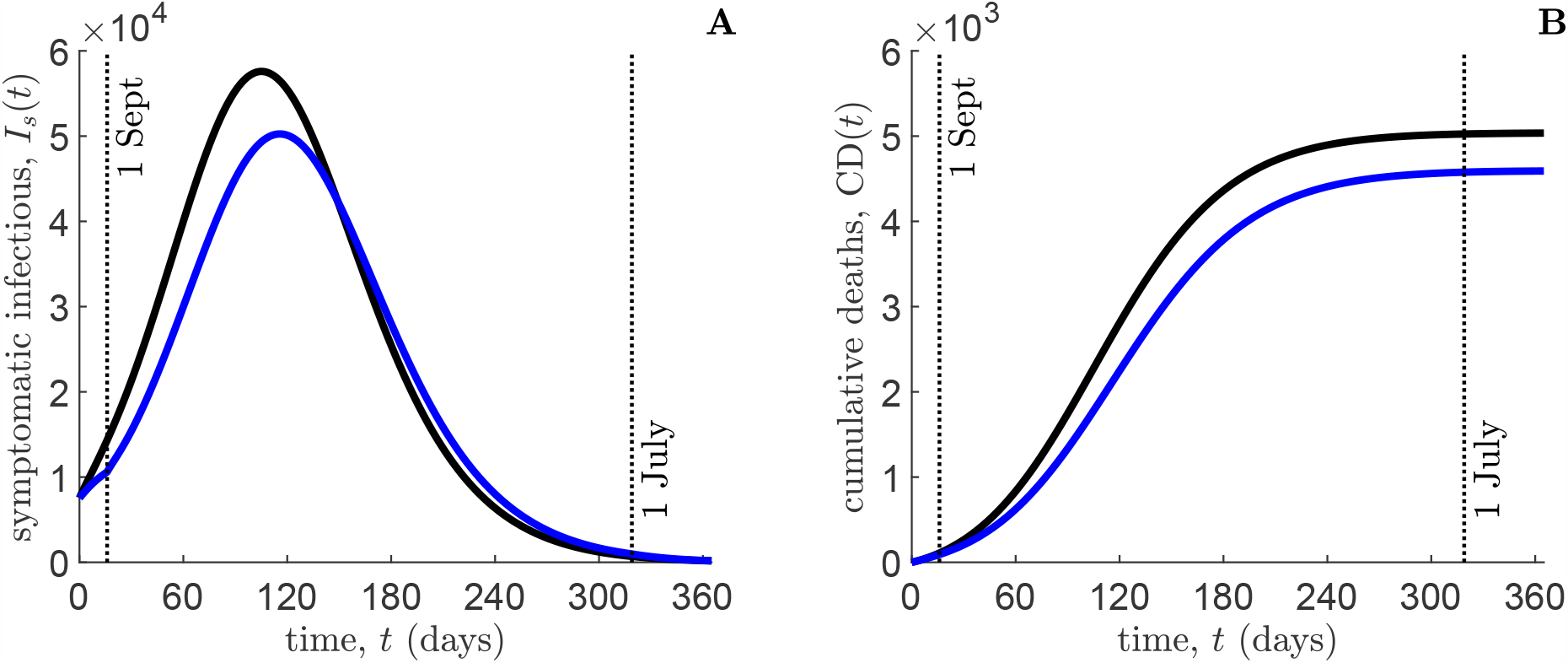
Impact of the seasonality on the information–dependent vaccination case (*φ*_0_ = 0.002 days^−1^, *D* = 500*μ/*Λ). Temporal dynamics of symptomatic infectious individuals *I*_*s*_ (panel A), and cumulative deaths CD(*t*) (panel B), as predicted by model (3)–(23). Blue lines: VAX-0S case (i.e. scenario including seasonality); black lines: VAX-0 case (i.e. no seasonality scenario). Initial conditions and other parameter values are given in Table 1 and in Section 6.

## 7 Conclusions

In this paper we introduced a mathematical model describing the transmission of the COVID–19 disease in presence of non mandatory vaccination. The main novelty is that the hesitancy and refusal of vaccination is taken into account. To this aim, we used the information index, which mimics the idea that individuals take their decision on vaccination based not only on the present but also on the past information they have on the spread of the disease.

Theoretical analysis and simulations show clearly as a voluntary vaccination can of course reduce the impact of the disease but it is unable to eliminate it. The qualitative path of the disease remains the same but the quantitative results are strongly different: an epidemic outbreak (a new epidemic wave) occurs, even if (as we observed in our simulations) the information-dependent vaccination rate is, at its peak, more than three times larger than the constant baseline vaccination rate.

A key result is in particular the fact that the information-related parameters deeply affect the dynamics of the disease: large information coverage and small memory characteristic time are needed to have the best results. The different impact of behaviour and information with respect to the scenario of mandatory constant vaccination can be further appreciated by examining the contour plots in Figs. 6-8.

As it is reasonable, the parameter *σ*, i.e. the risk of infection for vaccinated people, has a major impact.

Namely, the control reproduction number ℛ_*V*_ (*σ, φ*_0_) essentially depends on *σ* in a linear-affine manner. This suggest to stick to vaccines that have very low *σ*, where ℛ_*V*_ (*σ, φ*_0_) is tiny. A very positive result is that the threshold of non-efficacy of the vaccine, which can roughly be delineated as the curve (*σ, φ*_0_) where ℛ_*V*_ (*σ, φ*_0_) = 1 is located for values *σ* ∈ (0.6, 0.7), i.e. for very large values of *σ* (Figure 3A).

As far as the impact of human behaviour w.r.t. scenarios with constant vaccination rates is concerned, we obtained that that the performances were better only w.r.t. a constant vaccination rate as low as *φ*_0_, whereas the scenario where 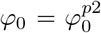(see Section 5.1) would lead to excellent result and a substantially smaller number of deaths.

As far as the comparison of the VAX-0 vs VAX-30 scenarios is concerned, we also measured its impact on the ratio between the information-dependent and the constant components of the vaccination rate, namely *φ*_1_(*M*)*/φ*_0_. As expected, the peak was considerably larger in the scenario VAX-30. The peaks occur in the same week if measured in the absolute time, i.e. the peak for VAX-30 occurs one month before the peaks of VAX-0 if measured in *time since the start of the vaccination*(see Figure 5).

Finally, seasonality has a relative but non neglectable relevance. For example, although the decrease of the transmission rate and of the onset of symptoms occur in the summer, the predicted winter epidemic peak of symptomatic cases is decreased and delayed w.r.t. the one in the no-seasonality scenario. A small but not neglectable decrease and delay of the cumulative deaths is also observed. This overall suggests that a decrease of the transmission and of the onset of symptoms has positive impact even many months after their end (see Figure 9).

An apparent limitation of this study is the absence of modelling for the dynamics of the transmission rate. In other words, neither spontaneous changes of the parameter *β* and imposed changes due to social distancing laws and partial/full lockdowns are taken into the account. However, these aspects are intentionally neglected here since our goal is to assess the impact of a possible voluntary vaccination campaign.

As far as future research is concerned, we plan: i) to explore (mainly numerically) a realistic model of the COVID-19 spread that includes the time-changes of the transmission rate; ii) to explore the possibility that eradication of the COVID-19 is not reached and the disease stays endemic.

## Data Availability

there are no external datasets or supplementary material online for this manuscript

## Acknowledgments

The present work has been performed under the auspices of the Italian National Group for the Mathematical Physics (GNFM) of National Institute for Advanced Mathematics (INdAM). M.G. thanks the support by the Italian National Research Project *Multiscale phenomena in Continuum Mechanics: singular limits; off-equilibrium and transitions* (PRIN 2017YBKNCE).

## A Alternative proof of Theorem 3

Consider the following function

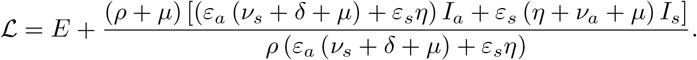

It is easily seen that the *ℒ* is non-negative in *𝒟* (see (6)) and also *ℒ* = 0 if and only if *E* = *I*_*a*_ = *I*_*s*_ = 0. The time derivative of *ℒ* along the solutions of system (3) in *𝒟* reads

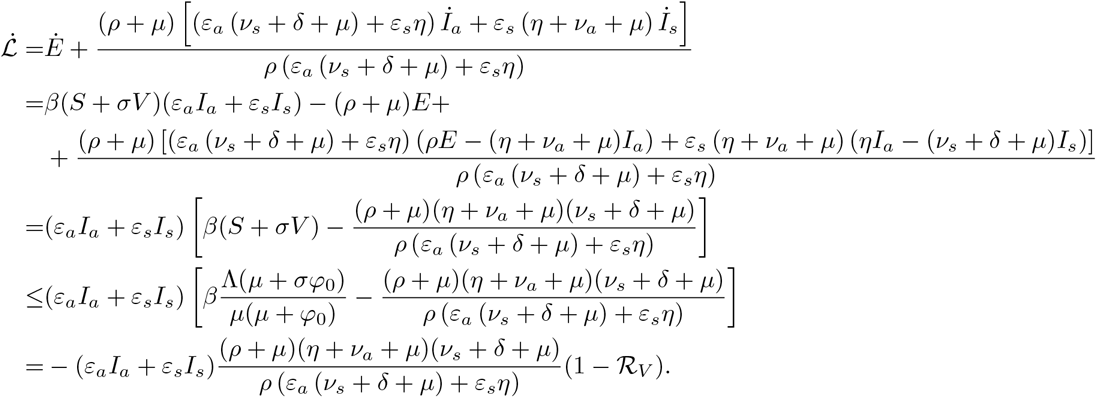

It follows that 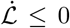 for ℛ_*V*_ < 1 with 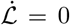 only if *I*_*a*_ = *I*_*s*_ = 0. Hence, ℒ is a Lyapunov function 𝒟 on and the largest compact invariant set in 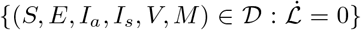 is the singleton DFE. Therefore, from the La Salle’s invariance principle [47], every solution to system (3) with initial conditions (4) approaches the DFE, as *t* → +∞.

